# Gender differences in the prevalence of Parkinson’s disease

**DOI:** 10.1101/2022.05.17.22275213

**Authors:** Alexandra Zirra, Shilpa C Rao, Jonathan Bestwick, Rajasumi Rajalingam, Connie Marras, Cornelis Blauwendraat, Ignacio Mata, Alastair J Noyce

## Abstract

**Background:** It is generally recognized that Parkinson’s disease (PD) affects males more commonly than females. The reasons for the difference in PD prevalence by gender remain unclear.

**Methods:** In this systematic review and meta-analysis, we updated previous work by searching MEDLINE, SCOPUS, and OVID for articles reporting PD prevalence for both genders between 2011-2021. We calculated overall male/female prevalence ratios (OPR) and investigated heterogeneity in effect estimates.

**Results:** 19 new and 13 previous articles were included. The OPR was 1.18, 95% CI [1.03, 1.36]. The OPR was lowest in Asia and appeared to be decreasing over time. Study design, national wealth, and participant age did not explain heterogeneity in OPR.

**Conclusion:** Gender differences in PD prevalence may not be as stark as previously thought, but still remain. Studies are needed to understand the role of genetic, environmental, and societal determinants of gender differences in prevalence.

## Introduction

The burden of Parkinson’s disease (PD) appears to be increasing and is estimated to reach 13 million PD cases by 2040.^1^ In observational studies, PD tends to affect males more frequently than females but the reasons are unclear.^2–5^ However, in some parts of the world (e.g. South Korea, Japan, Bolivia), a male predominance of PD is less apparent.^6–8^ Variation in prevalence may be explained by case ascertainment (specialist access), social determinants (health beliefs, insurance), or risk factors (i.e. genetic, environmental factors, or their interaction).^9–11^ Genetic factors are likely to be more prominent in patients with young-onset PD, defined by the age of onset <40 years. Male/female incidence and prevalence ratios for young-onset PD tend to be closer to 1,^1,12,13^ and sex-stratified genome-wide association studies (GWAS) have not found significant differences between genders.^14^

Pringsheim *et al* demonstrated that the gender ratio for prevalence increased with advancing age and varied by continent (lowest in Asian countries).^3^ In this systematic review and meta-analysis, we updated the knowledge about gender differences in PD prevalence, building on previous literature.^3^ Additionally, we sought out to understand whether the study design, economic profile of the country, age at inclusion, or life expectancy could explain gender differences in PD prevalence.

## Methods

### Data selection

In this systematic review and meta-analysis, we searched for studies published from 2011-2021 with the terms: “Parkinson” and “prevalence”/”epidemiology” on MEDLINE, SCOPUS, and OVID (**Table S1**). Articles were included if they were population-based (door-to-door surveys and healthcare registry studies using PD diagnostic codes/medication), reported prevalence rates for males and females separately, and data were stratified by age groups. We excluded cohort studies because their design captures incidence typically, and case-control studies due to potential selection bias.^15^ Other exclusion criteria were: lack of gender/age subgroup analysis, inclusion of secondary parkinsonism, and unavailable full-text.

Two independent reviewers (A.Z., S.C.R.) conducted the search and screened abstracts for eligibility. When differentially included, a third reviewer (A.J.N.) was involved. The list of selected prevalence studies was then combined with the list of studies in Pringsheim *et al*.^3^

### Data extraction

Extraction of prevalence data was performed by each reviewer including study reference, study design, target population, male/female prevalence, case numbers, total population by gender, and age group. Age-standardized or crude prevalence rates were used and converted to cases per 100,000 persons (**Supplementary material**).

Additional data were extracted from WHO life expectancy (LE) at birth from the year 2000 for studies published before this year and from the year 2010 for later studies.^16^ The LE gap was calculated as the difference between gender (female LE minus male LE). Age at inclusion was extracted from articles either as reported (median) or was calculated from frequency tables (as the rank of the median).^17^ World Bank data was used for the classification of high-income (HIC) and low/middle-income countries (L/MIC)^18^.

### Data analysis

Prevalence from each study was converted into a male/female ratio: OPR = overall male/female prevalence ratio with log transformation for normalization. A random-effects model was used due to high heterogeneity calculated by Cochrane Q statistic and I^2^. Subgroup meta-analysis and meta-regression were used to determine the significance of the different variables: study design, economic country profile, median age at study inclusion, LE gap, and study origin. For study origin Europe was used as the reference continent. STATA v.19 was used for statistical analysis and figures.

## Results

Our search identified 17,335 abstracts (**Figure 1A**). Titles and abstracts were screened for relevance, and to exclude case-control and cohort studies. From the first round, 350 articles were deemed relevant with 25 articles included for a full-text review after further exclusions. An in-depth review of the articles resulted in the exclusion of 6 articles due to insufficient data or no cases of PD identified for one gender (prevalence of 0). The final 19 articles from our search were added to the 13 articles from Pringsheim *et al* (**Figure 1B, Table S2, Supplementary References**).^3^

**Figure 1.**
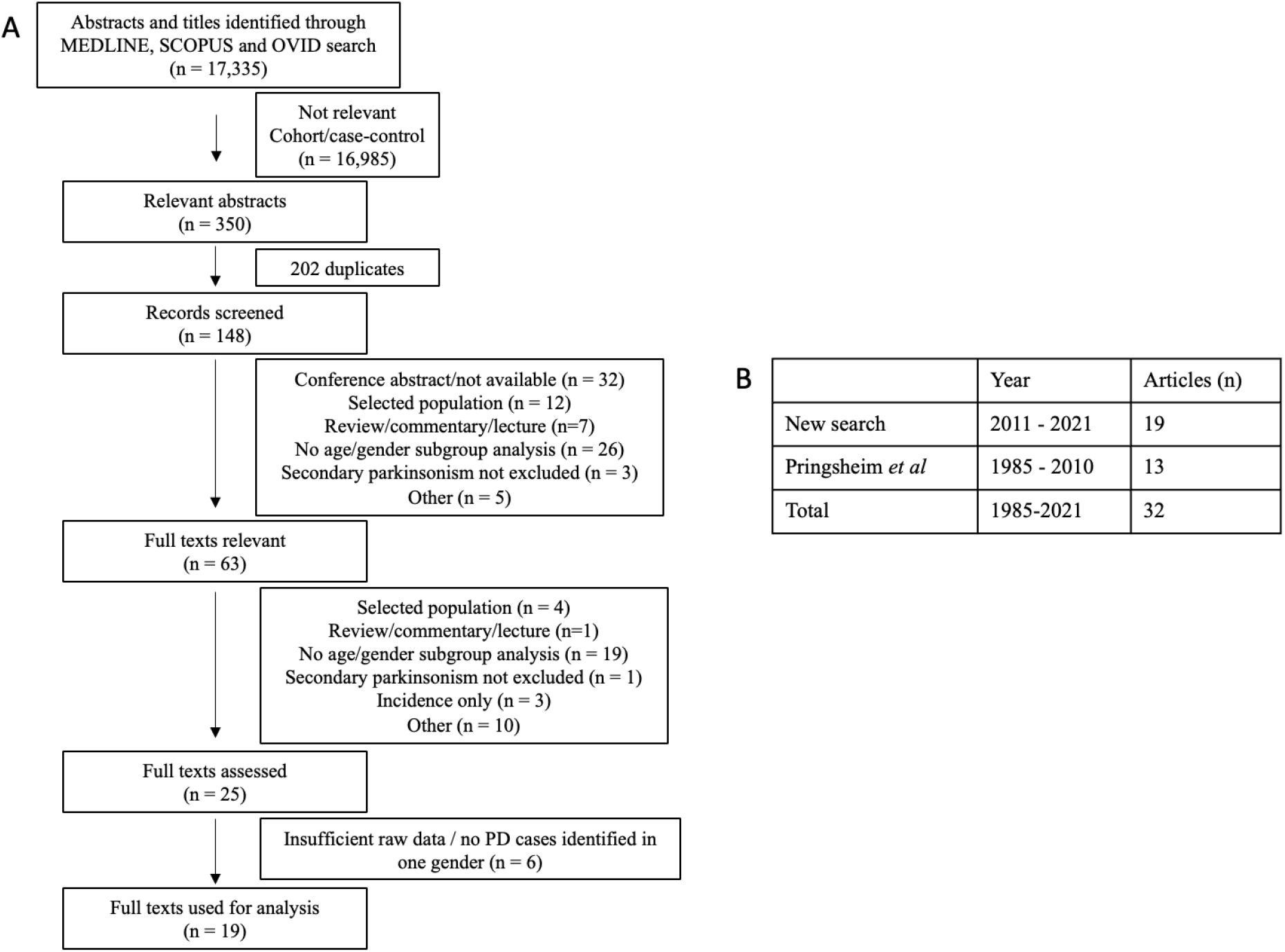
PRISMA Flow diagram of article selection and final count of articles selected. Table 1: (A) Results from search strategy (B) Final results from search strategy included a total of 19 articles from 2011 to 2021. These were added to the list of articles from Prinsheim et al., totaling 32 articles from the years 1985 to 2021.

From the pooling effect estimates from a random-effects model of the 32 articles, we calculated an OPR of 1.18, 95% CI [1.03,1.36] with a high degree of heterogeneity (I^2^=99.8%) (**Figure S1A**). There was no evidence of small study bias (p = 0.562; **Figure S1B**).

We explored the high heterogeneity observed in the data by considering study design (door-to-door survey versus health records analysis), national economic wealth (HIC versus L/MIC), and continent. Meta-regression for study design did not provide evidence that this was a source of heterogeneity (p = 0.141). However, studies from L/MIC had a higher OPR of 1.28, 95% CI [1.16,1.4] compared to studies from HIC of 1.14, 95% CI [0.95,1.35]. When we looked at OPR between continents, Asia was found to have the smallest OPR, 1.04, 95% CI [0.83,1.29] (**Figure 2**). Meta-regression for categorical data - economic performance and continent, did not provide evidence that either were sources of heterogeneity (p = 0.362 and p = 0.100 for Asian compared to European studies, respectively).

**Figure 2.**
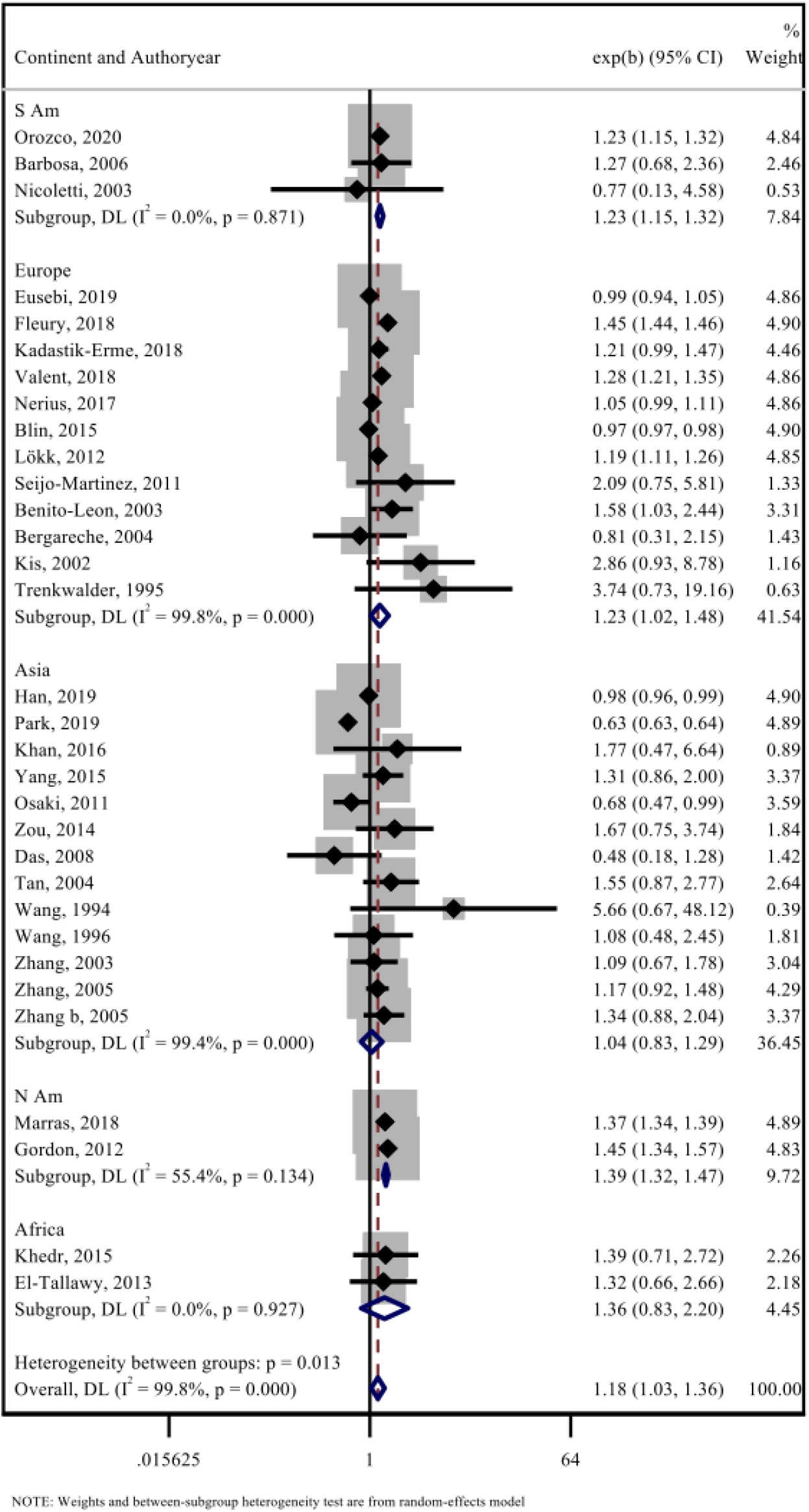
Subgroup meta-analysis using random effects model for each study continent in the M/F prevalence ratio. Random effects meta-analysis with Cochrane Q statistic of the male/female prevalence ratio of each study categorized by the study’s respective continent. The pooled prevalence ratio for all the studies is 1.18, 95% CI [1.03,1.36], but heterogeneity is high between groups (I^2^= 99.8%, *p* = 0.013).

Next, we investigated whether age at inclusion or LE gap could explain the heterogeneity. The median age for PD patients from the 32 studies was 75.4 (range 55-85). Meta-regression using median age did not suggest that this was a source of observed heterogeneity (p = 0.059). The LE gap also did not account for observed heterogeneity (p = 0.080).

Lastly, we investigated whether OPR varied with publication year. A subgroup analysis showed a decreasing trend in the last 30 years: 1990-2000 OPR 2.06, 95% CI [0.72, 5.87], 2000-2010 OPR 1.24, 95% CI [1.06,1.45] and 2010-present OPR 1.15, 95% CI[0.98,1.35], p = 0.019 (**Figure S2**).

## Discussion

This systematic review and meta-analysis demonstrated that the male/female prevalence ratio for PD is lower than has previously been reported by other studies.^2^ The lowest male/female ratio was seen in studies from Asia. We also observed that OPR may be decreasing over time and that it was higher in L/MIC compared with HIC.

Large population-based studies are best suited for determining the prevalence of diseases like PD.^15^ We included data from healthcare registries and door-to-door cross-sectional studies. Healthcare registries may cover 50-100% of a particular population,^6,19^ thus for population-based prevalence estimates, door-to-door studies are considered the gold standard. PD diagnostic criteria vary across regions and institutions, causing potential differences in prevalence estimates for door-to-door and healthcare registries.^19,20^ Our analysis did not identify study design as a source of heterogeneity in prevalence ratios.

There was little evidence to suggest that age at inclusion (AAO) might explain differences in gender ratio. Previous studies have suggested that in patients with younger AAO, the ratio is closer to 1.^2^ One might expect that a younger AAO may arise to a greater contribution from genetic factors. GWAS studies to date, have not identified different genetic risk factors for PD between sexes,^14^ but these studies over-represent participants of European ancestry and have lower female inclusion rates.^9^ Efforts to create large trans-ethnic genomic datasets will help us to better understand PD genetics in under-represented populations.^21^

A source of heterogeneity that we were unable to explore properly was the role of environmental factors. Rural living and occupational pesticide exposure are associated with an increase in PD risk.^22,23^ Males have traditionally held agricultural occupations, which could explain the higher male PD prevalence.^24^ Increasing urbanization decreases in agriculture, and organophosphate use in some regions may explain a diminishing OPR. Other putative risks/protective factors, including the Mediterranean diet, type 2 diabetes, smoking, and alcohol consumption,^25–30^ might contribute to the male predominance of PD.

Healthcare inequalities could account for observed differences in male/female PD prevalence. In L/MIC, such as Benin, 60% of females reported socioeconomic and marital barriers to accessing healthcare.^31,32^ Stigma related to PD symptoms and cultural beliefs about ‘normal’ aging/PD awareness may explain the different levels of healthcare access globally.^33^ Our data showed that studies from L/MIC have a higher OPR, which may reflect unequal access to healthcare between genders.

Another important consideration is the relationship between disease prevalence, incidence, and survival. Females have a higher AAO and a lower incidence of PD but also have a longer LE than males.^34^ Female longevity may influence the male/female prevalence imbalance in other neurodegenerative diseases, like Alzheimer’s.^35^ Our analysis did not support this hypothesis as life expectancy did not explain heterogeneity in OPR. However, the LE gap has changed over time,^36^ and this may partly explain why the OPR is also seen to be decreasing in the last 30 years. The recent decrease in life expectancy gap may be related to environmental exposure, such as increased smoking rates in the female population^37^ or the increasing integration of females into jobs traditionally held by males. Further studies are needed to understand the effect of LE and environmental/societal factors on PD prevalence.

A limitation of our study was not conducting the search prior to 2011. Consequently, we included both door-to-door studies - similar to Pringsheim *et al*, as well as studies with the newer methodology of predictive algorithms. However, no significant difference was detected between the newly identified studies and those from Pringsheim *et al*.^3^ Another limitation in our study was the use of both crude and age-standardized prevalence ratios. A third limitation was classifying the articles by geographic region since PD is a multifactorial disease in which genetics, environmental, and demographic factors may vary even between countries from the same region.^38^ This may have impacted our results and further studies of differences within regions are needed.

In conclusion, this systematic review and meta-analysis shows that the gender ratio for PD prevalence is lower in Asian populations and may have been decreasing in recent years. Further high-quality epidemiological studies of diverse populations are needed to understand whether the effect stems from environmental, societal, and/or genetic factors.

## Supporting information

Raw data

Code repository

## Data Availability

All data produced in the present work are contained in the manuscript and supplementary files.

## Supplementary

Raw data: “Prevalence v5.pdf”.

Code repository: “Prevalence MA code.pdf”.

**Supplementary Table S1.** Search strategies for MEDLINE, SCOPUS and OVID.

| DATABASE | SEARCH STRATEGY |
| --- | --- |
| MEDLINE | "Parkinson disease"[All Fields] AND "prevalence"[All Fields]; Filters: Journal Article, English, MEDLINE, from 2011/1/1 - 2021/12/30 Sort by: Most Recent |
|  | "Parkinson disease"[All Fields] AND "epidemiology"[All Fields]; Filters: Journal Article, English, MEDLINE, from 2011/1/1 - 2021/12/30 Sort by: Most Recent |
| SCOPUS | ( TITLE-ABS-KEY ( {Parkinson disease} AND {prevalence} ) AND LANGUAGE ( English ) ) AND PUBYEAR > 2011 |
|  | ( TITLE-ABS-KEY ( {Parkinson disease} AND {epidemiology} ) AND LANGUAGE ( English ) ) AND PUBYEAR > 2011 |
| OVID | "Parkinson disease" AND "prevalence" {Including Limited Related Terms}; Ovid, 2011 to current, English |
|  | "Parkinson disease" AND "epidemiology" {Including Limited Related Terms}; Ovid, 2011 to current, English |

**Table S2.** List of included prevalence studies with study characteristics.

| Author, year | OPR | CI |  | Age* | M life exp | F life exp | F-M survival difference | Age group | Study type | Continent |
| --- | --- | --- | --- | --- | --- | --- | --- | --- | --- | --- |
| Orozco, 2020 | 1.23 | 1.15 | 1.32 | 73 | 74 | 80.15 | 6.15 | 30+ | P | S America |
| Eusebi, 2019 | 0.99 | 0.94 | 1.05 | 82 | 79.47 | 84.22 | 4.75 | 40+ | P | Europe |
| Han, 2019 | 0.98 | 0.96 | 0.99 | 72 | 77.12 | 83.78 | 6.66 | 0+ | P | Asia |
| Park, 2019 | 0.63 | 0.63 | 0.64 | 85 | 77.12 | 83.78 | 6.66 | 0+ | P | Asia |
| Fleury, 2018 | 1.45 | 1.44 | 1.46 | 75 | 80.03 | 84.18 | 4.15 | 0+ | P | Europe |
| Kadastik-Erme, 2018 | 1.21 | 0.99 | 1.47 | 77 | 70.86 | 80.45 | 9.59 | 0+ | P | Europe |
| Marras, 2018 | 1.37 | 1.34 | 1.39 | 80 | 78.56 | 80.8 | 2.24 | 45+ | P | N America |
| Valent, 2018 | 1.28 | 1.21 | 1.35 | 77 | 79.47 | 84.22 | 4.75 | 0+ | P | Europe |
| Nerius, 2017 | 1.05 | 0.99 | 1.11 | 77 | 77.78 | 82.54 | 4.76 | 0+ | P | Europe |
| Khan, 2016 | 1.77 | 0.47 | 6.64 | 72 | 61.53 | 63.9 | 2.37 | 50+ | S | Asia |
| Blin, 2015 | 0.97 | 0.97 | 0.98 | 80 | 77.96 | 84.34 | 6.38 | 18+ | P | Europe |
| Khedr, 2015 | 1.39 | 0.71 | 2.72 | 67 | 67.82 | 72.69 | 4.87 | 0+ | S | Africa |
| Yang, 2015 | 1.31 | 0.86 | 2.00 | 70 | 72.26 | 77.94 | 5.68 | 0+ | S | Asia |
| Zou, 2014 | 1.67 | 0.75 | 3.74 | 85 | 72.26 | 77.94 | 5.68 | 70+ | P | Asia |
| El-Tallawy, 2013 | 1.32 | 0.66 | 2.66 | 65 | 67.82 | 72.69 | 4.87 | 40+ | S | Africa |
| Gordon, 2012 | 1.45 | 1.34 | 1.57 | 70 | 78.56 | 80.8 | 2.24 | 40+ | P | N America |
| Lökk, 2012 | 1.19 | 1.11 | 1.26 | 74 | 79.41 | 83.15 | 3.74 | 0+ | P | Europe |
| Seijo-Martinez, 2011 | 2.09 | 0.75 | 5.81 | 80 | 78.76 | 84.58 | 5.82 | 65+ | S | Europe |
| Osaki, 2011 | 0.68 | 0.47 | 0.99 | 75 | 79.46 | 85.77 | 6.31 | 0+ | P | Asia |
| Das, 2008 | 0.48 | 0.18 | 1.28 | 80 | 65.69 | 68.94 | 3.25 | 60+ | S | India |
| Barbosa, 2006 | 1.27 | 0.68 | 2.36 | 82 | 70.57 | 77.98 | 7.41 | 65+ | S | S America |
| Zhang, 2005 | 1.17 | 0.92 | 1.48 | 80 | 72.26 | 77.94 | 5.68 | 55+ | S | Asia |
| Zhang b, 2005 | 1.34 | 0.88 | 2.04 | 63 | 72.26 | 77.94 | 5.68 | 50+ | S | Asia |
| Bergareche, 2004 | 0.81 | 0.31 | 2.15 | 80 | 78.76 | 84.58 | 5.82 | 65+ | S | Europe |
| Tan, 2004 | 1.55 | 0.87 | 2.77 | 75 | 79.36 | 84.01 | 4.65 | 50+ | S | Asia |
| Benito-Leon, 2003 | 1.58 | 1.03 | 2.44 | 77 | 78.76 | 84.58 | 5.82 | 65+ | S | Europe |
| Nicoletti, 2003 | 0.77 | 0.13 | 4.58 | 55 | 79.47 | 84.22 | 4.75 | 40+ | S | Europe |
| Zhang, 2003 | 1.09 | 0.67 | 1.78 | 80 | 72.26 | 77.94 | 5.68 | 55+ | S | Asia |
| Kis, 2002 | 2.86 | 0.93 | 8.78 | 80 | 79.47 | 84.22 | 4.75 | 60+ | S | Europe |
| Wang, 1996 | 1.08 | 0.48 | 2.45 | 71.3 | 69.34 | 74.15 | 4.81 | 50+ | S | Asia |
| Trenkwalder, 1995 | 3.74 | 0.73 | 19.16 | 78 | 75.04 | 80.92 | 5.88 | 65+ | S | Europe |
| Wang, 1994 | 5.66 | 0.67 | 48.12 | 76.5 | 69.34 | 74.15 | 4.81 | 50+ | S | Asia |

OPR – overall male to female prevalence ratio; CI – confidence interval; exp – expectancy; F – female; M – male; P – predictive, S – survey. \* age of inclusion is recorded as median. Life expectancy values for each gender were extracted from WHO life expectancy (LE) at birth from the year 2000 for studies published before this year and from the year 2010 for later studies.

**Supplementary Figure S1.**
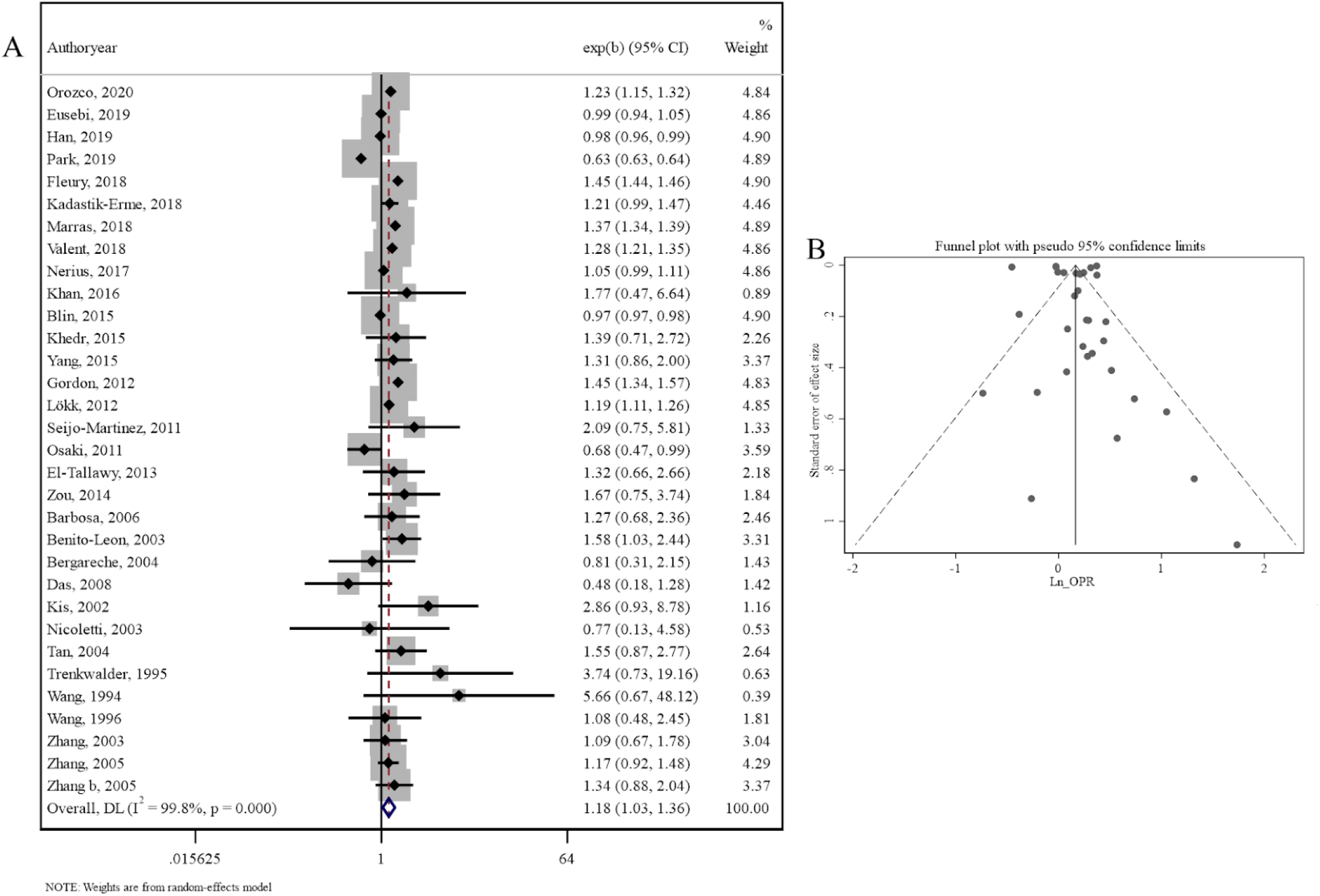
Meta-funnel plot analyses for reporting bias in prevalence studies. (A) Meta-analysis using random effects and Cochrane Q statistic for male/female prevalence ratios with an overall prevalence ratio of 1.18, 95% CI [1.03,1.36]. (B) Metafunnel plot of included studies did not identify any evidence of small studying reporting bias (p = 0.562).

**Supplementary Figure S2.**
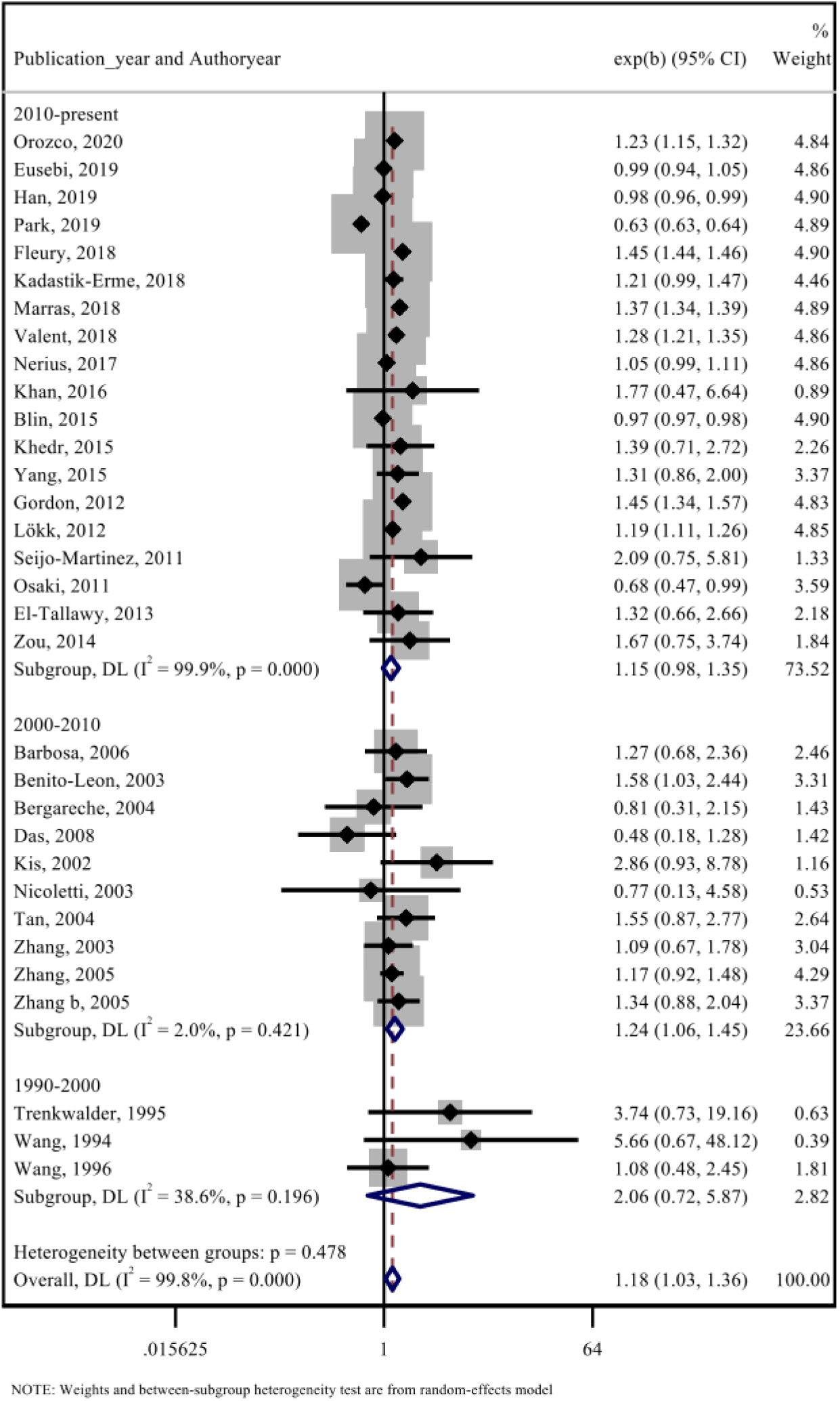
Subgroup meta-analysis using random effects model for time-trends in the M/F prevalence ratio. Random effects meta-analysis with Cochrane Q statistic of the M/F prevalence prevalence ratio of each study categorized by the year of publication. This shows a decreasing trend from 2.06, 95% CI [0.72, 5.87] in 1990-2000 to 1.24, 95% CI [1.06, 1.45] in 2000-2010, and to 1.15, 95% CI [0.98, 1.35] in 2010-2021.

## Notes

**Relevant conflicts of interest/financial disclosures:** Nothing to report.

### Competing Interest Statement

A.J.N. reports grants from the Barts Charity, Parkinson’s UK, Aligning Science Across Parkinson’s and Michael J. Fox Foundation, and the Virginia Keiley Benefaction. Personal fees/honoraria from Britannia, BIAL, AbbVie, Global Kinetics Corporation, Profile, Biogen, Roche, and UCB are outside of the submitted work. I.F.M. reports grants from Aligning Science Across Parkinson’s and Michael J. Fox Foundation, The Parkinsons Foundation, American Parkinson Disease Association, and National Institutes of Health.

### Author Declarations

We have used published articles on prevalence of PD in both genders. The references are available in the file "Prevalence v5.pdf" (see supplementary material).

