## Supplementary material for "Gender differences in the prevalence of Parkinson’s disease": Raw data

| Author | Year | Authoryear | Prevalence_M | PD_M | Total_M | NonPD_M | Prevalence_F | PD_F | Total_F | NonPD_F | Total_cases | Total_pop | OPR | Ln_OPR | Lower_CI_95 | Upper_CI_95 | Ln_lower_CI_95 | Ln_upper_CI_95 | Age_group | Country_reclass | Continent | Study_type | Median_age | Life_expectancy_M | Life_expectancy_F | F_to_M_difference_in_Life_expectancy | Publication_year | Update |
|---|---|---|---|---|---|---|---|---|---|---|---|---|---|---|---|---|---|---|---|---|---|---|---|---|---|---|---|---|
| Orozco | 2020 | Orozco, 2020 | 175 | 1750 | 1000000 | 998250 | 141.91 | 1514 | 1066873 | 1065359 | 3264 | 2066873 | 1.2332 | 0.2096 | 1.1513 | 1.3209 | 0.1409 | 0.2783 | 0 | 2 | S Am | 2 | 73 | 74 | 80.15 | 6.15 | 2010-present | 1 |
| Eusebi | 2019 | Eusebi, 2019 | 540 | 2628 | 486667 | 484039 | 544 | 2848 | 523529 | 520681 | 5476 | 1010196 | 0.9926 | -0.0074 | 0.9415 | 1.0465 | -0.0603 | 0.0455 | 2 | 1 | Europe | 2 | 82 | 79.47 | 84.22 | 4.75 | 2010-present | 1 |
| Han | 2019 | Han, 2019 | 113.1 | 38157 | 33737401 | 33699244 | 116 | 54236 | 46755172 | 46700936 | 92393 | 80492573 | 0.975 | -0.0253 | 0.9623 | 0.9878 | -0.0384 | -0.0122 | 0 | 1 | Asia | 2 | 72 | 77.12 | 83.78 | 6.66 | 2010-present | 1 |
| Park | 2019 | Park, 2019 | 108.4 | 32002 | 29522140 | 29490138 | 170.8 | 48745 | 28539227 | 28490482 | 80747 | 58061367 | 0.6347 | -0.4547 | 0.6258 | 0.6437 | -0.4688 | -0.4406 | 0 | 1 | Asia | 2 | 85 | 77.12 | 83.78 | 6.66 | 2010-present | 1 |
| Fleury | 2018 | Fleury, 2018 | 223.7 | 227697 | 101786768 | 101559071 | 154.5 | 242815 | 157161812 | 156918997 | 470512 | 258948580 | 1.4479 | 0.3701 | 1.4396 | 1.4562 | 0.3644 | 0.3758 | 0 | 1 | Europe | 2 | 75 | 80.03 | 84.18 | 4.15 | 2010-present | 1 |
| Kadastik-Erme | 2018 | Kadastik-Erme, 2018 | 355 | 160 | 45070 | 44910 | 294 | 271 | 92177 | 91906 | 431 | 137247 | 1.2075 | 0.1885 | 0.9935 | 1.4676 | -0.0065 | 0.3836 | 0 | 1 | Europe | 2 | 77 | 70.86 | 80.45 | 9.59 | 2010-present | 1 |
| Marras | 2018 | Marras, 2018 | 667 | 20366 | 3053373 | 3033007 | 488 | 23491 | 4813730 | 4790239 | 43857 | 7867103 | 1.3668 | 0.3125 | 1.3415 | 1.3926 | 0.2938 | 0.3312 | 3 | 1 | N Am | 2 | 80 | 78.56 | 80.8 | 2.24 | 2010-present | 1 |
| Valent | 2018 | Valent, 2018 | 747.08 | 2270 | 303850 | 301580 | 585.33 | 2465 | 421130 | 418665 | 4735 | 724980 | 1.2763 | 0.244 | 1.2058 | 1.351 | 0.1872 | 0.3008 | 0 | 1 | Europe | 2 | 77 | 79.47 | 84.22 | 4.75 | 2010-present | 1 |
| Nerius | 2017 | Nerius, 2017 | 820 | 2002 | 244146 | 242144 | 779 | 2734 | 350963 | 348229 | 4736 | 595109 | 1.0526 | 0.0513 | 0.9939 | 1.1148 | -0.0061 | 0.1087 | 0 | 1 | Europe | 2 | 77 | 77.78 | 82.54 | 4.76 | 2010-present | 1 |
| Khan | 2016 | Khan, 2016 | 530 | 8 | 1509 | 1501 | 300 | 3 | 1000 | 997 | 11 | 2509 | 1.7667 | 0.5691 | 0.4698 | 6.6433 | -0.7554 | 1.8936 | 4 | 2 | Asia | 1 | 72 | 61.53 | 63.9 | 2.37 | 2010-present | 1 |
| Blin | 2015 | Blin, 2015 | 304 | 95716 | 31485526 | 31389810 | 312 | 104557 | 33511859 | 33407302 | 200273 | 64997385 | 0.9744 | -0.026 | 0.9659 | 0.9829 | -0.0347 | -0.0172 | 1 | 1 | Europe | 2 | 80 | 77.96 | 84.34 | 6.38 | 2010-present | 1 |
| Khedr | 2015 | Khedr, 2015 | 503 | 21 | 4175 | 4154 | 363 | 14 | 3857 | 3843 | 35 | 8032 | 1.3857 | 0.3262 | 0.7056 | 2.7211 | -0.3487 | 1.001 | 0 | 2 | Africa | 1 | 67 | 67.82 | 72.69 | 4.87 | 2010-present | 1 |
| Yang | 2015 | Yang, 2015 | 1680 | 51 | 3036 | 2985 | 1280 | 37 | 2891 | 2854 | 88 | 5926 | 1.3125 | 0.2719 | 0.8622 | 1.998 | -0.1483 | 0.6921 | 0 | 2 | Asia | 1 | 70 | 72.26 | 77.94 | 5.68 | 2010-present | 1 |
| Zou | 2014 | Zou, 2014 | 2440 | 219 | 8975 | 8756 | 1460 | 6 | 411 | 405 | 225 | 9386 | 1.6712 | 0.5136 | 0.7472 | 3.7381 | -0.2914 | 1.3186 | 8 | 1 | Asia | 2 | 85 | 72.26 | 77.94 | 5.68 | 2010-present | 1 |
| El-Tallawy | 2013 | El-Tallawy, 2013 | 240.38 | 20 | 8320 | 8300 | 181.51 | 13 | 7162 | 7149 | 33 | 15482 | 1.3243 | 0.2809 | 0.6593 | 2.6604 | -0.4166 | 0.9785 | 2 | 1 | Africa | 1 | 65 | 67.82 | 72.69 | 4.87 | 2010-present | 1 |
| Gordon | 2012 | Gordon, 2012 | 1012 | 1322 | 130632 | 129310 | 697.2 | 1291 | 185169 | 183878 | 2613 | 315802 | 1.4515 | 0.3726 | 1.3448 | 1.5667 | 0.2962 | 0.449 | 2 | 2 | N Am | 2 | 70 | 78.56 | 80.8 | 2.24 | 2010-present | 1 |
| Lökk | 2012 | Lökk, 2012 | 213.18 | 2127 | 997748 | 995621 | 179.74 | 1836 | 1021475 | 1019639 | 3963 | 2019224 | 1.186 | 0.1706 | 1.1143 | 1.2624 | 0.1082 | 0.233 | 0 | 2 | Europe | 2 | 74 | 79.41 | 83.15 | 3.74 | 2010-present | 1 |
| Seijo-Martinez | 2011 | Seijo-Martinez, 2011 | 2860 | 9 | 315 | 306 | 1370 | 6 | 438 | 432 | 15 | 753 | 2.0876 | 0.736 | 0.7507 | 5.8056 | -0.2868 | 1.7588 | 7 | 1 | Europe | 1 | 80 | 78.76 | 84.58 | 5.82 | 2010-present | 1 |
| Osaki | 2011 | Osaki, 2011 | 88 | 43 | 48864 | 48821 | 129 | 73 | 56589 | 56516 | 116 | 105453 | 0.6822 | -0.3825 | 0.4681 | 0.9941 | -0.7591 | -0.0059 | 0 | 1 | Asia | 2 | 75 | 79.46 | 85.77 | 6.31 | 2010-present | 1 |
| Das | 2008 | Das, 2008 | 216 | 6 | 2772 | 2766 | 451 | 12 | 2658 | 2646 | 18 | 5430 | 0.4789 | -0.7362 | 0.1799 | 1.2753 | -1.7156 | 0.2432 | 6 | 2 | Asia | 1 | 80 | 65.69 | 68.94 | 3.25 | 2000-2010 | 0 |
| Barbosa | 2006 | Barbosa, 2006 | 3800 | 17 | 447 | 430 | 3000 | 22 | 733 | 711 | 39 | 1181 | 1.2667 | 0.2364 | 0.6801 | 2.3591 | -0.3855 | 0.8583 | 7 | 2 | S Am | 1 | 82 | 70.57 | 77.98 | 7.41 | 2000-2010 | 0 |
| Zhang | 2005 | Zhang, 2005 | 1040 | 130 | 12500 | 12370 | 890 | 147 | 16517 | 16370 | 277 | 29017 | 1.1685 | 0.1558 | 0.924 | 1.4778 | -0.0791 | 0.3906 | 5 | 1 | Asia | 1 | 80 | 72.26 | 77.94 | 5.68 | 2000-2010 | 0 |
| Zhang b | 2005 | Zhang b, 2005 | 614 | 41 | 6678 | 6637 | 459 | 45 | 9804 | 9759 | 86 | 16481 | 1.3377 | 0.2909 | 0.8772 | 2.04 | -0.1311 | 0.713 | 4 | 2 | Asia | 1 | 63 | 72.26 | 77.94 | 5.68 | 2000-2010 | 0 |
| Bergareche | 2004 | Bergareche, 2004 | 1300 | 6 | 462 | 456 | 1600 | 12 | 750 | 738 | 18 | 1212 | 0.8125 | -0.2076 | 0.307 | 2.15 | -1.1808 | 0.7655 | 7 | 2 | Europe | 1 | 80 | 78.76 | 84.58 | 5.82 | 2000-2010 | 0 |
| Tan | 2004 | Tan, 2004 | 310 | 25 | 8065 | 8040 | 200 | 21 | 10500 | 10479 | 46 | 18565 | 1.55 | 0.4383 | 0.8683 | 2.7668 | -0.1412 | 1.0177 | 4 | 1 | Asia | 1 | 75 | 79.36 | 84.01 | 4.65 | 2000-2010 | 0 |
| Benito-Leon | 2003 | Benito-Leon, 2003 | 1900 | 43 | 2263 | 2220 | 1200 | 38 | 3167 | 3129 | 81 | 5430 | 1.5833 | 0.4595 | 1.0268 | 2.4414 | 0.0265 | 0.8926 | 7 | 2 | Europe | 1 | 77 | 78.76 | 84.58 | 5.82 | 2000-2010 | 0 |
| Nicoletti | 2003 | Nicoletti, 2003 | 248 | 2 | 806 | 804 | 323 | 3 | 929 | 926 | 5 | 1735 | 0.7678 | -0.2642 | 0.1286 | 4.5837 | -2.051 | 1.5225 | 2 | 1 | S Am | 1 | 55 | 79.47 | 84.22 | 4.75 | 2000-2010 | 0 |
| Zhang | 2003 | Zhang, 2003 | 1200 | 30 | 2500 | 2470 | 1100 | 34 | 3091 | 3057 | 64 | 5591 | 1.0909 | 0.087 | 0.6696 | 1.7774 | -0.4011 | 0.5751 | 5 | 2 | Asia | 1 | 80 | 72.26 | 77.94 | 5.68 | 2000-2010 | 0 |
| Kis | 2002 | Kis, 2002 | 4800 | 6 | 336 | 330 | 1680 | 6 | 414 | 408 | 12 | 750 | 2.8571 | 1.0498 | 0.93 | 8.7779 | -0.0726 | 2.1722 | 6 | 1 | Europe | 1 | 80 | 79.47 | 84.22 | 4.75 | 2000-2010 | 0 |
| Wang | 1996 | Wang, 1996 | 610 | 12 | 1967 | 1955 | 564 | 11 | 1950 | 1939 | 23 | 3918 | 1.0816 | 0.0784 | 0.4784 | 2.4453 | -0.7373 | 0.8942 | 4 | 1 | Asia | 1 | 71 | 69.34 | 74.15 | 4.81 | 1990-2000 | 0 |
| Trenkwalder | 1995 | Trenkwalder, 1995 | 1270 | 5 | 394 | 389 | 340 | 2 | 588 | 586 | 7 | 982 | 3.7353 | 1.3178 | 0.7283 | 19.1577 | -0.3171 | 2.9527 | 7 | 1 | Europe | 1 | 78 | 75.04 | 80.92 | 5.88 | 1990-2000 | 0 |
| Wang | 1994 | Wang, 1994 | 2212.39 | 5 | 226 | 221 | 390.63 | 1 | 256 | 255 | 6 | 482 | 5.6637 | 1.7341 | 0.6666 | 48.1195 | -0.4055 | 3.8737 | 4 | 2 | Asia | 1 | 77 | 69.34 | 74.15 | 4.81 | 1990-2000 | 0 |

| | | | | | | | | | | |
|---|---|---|---|---|---|---|---|---|---|---|
| Header | Explanation | | | | | | | | | |
| Author | First author | | | | | | | | | |
| Year | Year of publication | | | | | | | | | |
| Authoryear | | | | | | | | | | |
| Prevalence_M | Reported male prevalence / 100,000 persons | | | | | | | | | |
| PD_M | Number of male PD cases | | | | | | | | | |
| Total_M | Total number of males in population | | | | | | | | | |
| NonPD_M | Number of males without PD | | | | | | | | | |
| Prevalence_F | Reported female prevalence / 100,000 persons | | | | | | | | | |
| PD_F | Number of female PD cases | | | | | | | | | |
| Total_F | Total number of females in population | | | | | | | | | |
| NonPD_F | Number of females without PD | | | | | | | | | |
| Total_cases | Total number of PD cases in population | | | | | | | | | |
| Total_pop | Total population | | | | | | | | | |
| OPR | Overall prevalence rate (Male-to-Female) | | | | | | | | | |
| Ln_OPR | Natural logarithm of OPR | | | | | | | | | |
| Lower_CI_95 | 95% lower CI | | | | | | | | | |
| Upper_CI_95 | 95% upper CI | | | | | | | | | |
| Ln_lower_CI_95 | Natural logarithm of 95% lower CI | | | | | | | | | |
| Ln_upper_CI_95 | Natural logarithm of 95% upper CI | | | | | | | | | |
| Age_group | Ages groups included in study | 0 - all ages | 1 - 18+ | 2 - 40+ | 3 - 45+ | 4 - 50+ | 5 - 55+ | 6 - 60+ | 7 - 65+ | 8 - 70+ |
| Country_reclass | Economic profile of country | 1 - HIC | 2 - L/MIC | | | | | | | |
| Continent | Study continent | | | | | | | | | |
| Study_type | Study design | 1 - Door-to-door | 2 - Predictive | | | | | | | |
| Median_age | Median age at inclusion | | | | | | | | | |
| Life_expectancy_M | Male life expectancy at birth based on WHO data | | | | | | | | | |
| Life_expectancy_F | Female life expectancy at birth based on WHO data | | | | | | | | | |
| F_to_M_difference_in_Life_expectancy | Difference between female and male life expectancy | | | | | | | | | |
| Publication_year | Year of publication | 1990-2000 | 2000-2010 | 2010-present | | | | | | |
| Update | Article from new/old search | 1 - New search | 0 - Pringsheim *et al* | | | | | | | |
