## Supplementary material for "Gender differences in the prevalence of Parkinson’s disease": Code repository

**PREVALENCE**

**#Set working directory and import csv file**

cd "H:\Gender MA"

import delimited "H:\Gender MA\Data\Prevalence v4.csv"

**#Install functions**

ssc install metan

ssc install metareg

ssc install metabias

ssc install metafunnel

**#Run MA:**

meta set ln_opr ln_lower_ci_95 ln_upper_ci_95, studylabel(authoryear) random civartolerance(0.01)

metan ln_opr ln_lower_ci_95 ln_upper_ci_95, random eform label(namevar= authoryear)

Overall RR: 1.181	(1.028 ; 1.356); Heterogeneity I^2: 99.8%

**#Run publication bias analysis:**

metabias ln_opr _seES, egger

metafunnel ln_opr _seES

There is no significant publication bias, p = 0.562 (metabias)

**#Run meta-regression for binary variables:**

**First analysis**

xi: metareg ln_opr i.country, wsse(_seES )

High vs low income does not explain the heterogeneity p = 0.2 (metareg**)**
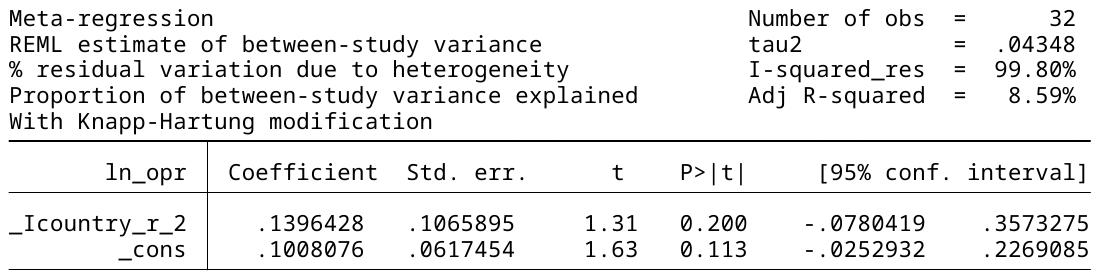

**Second analysis** – with updated HIC vs non-HIC

xi: metareg ln_opr i.country_reclass , wsse(_seES )

Second analysis: no difference, p = 0.200

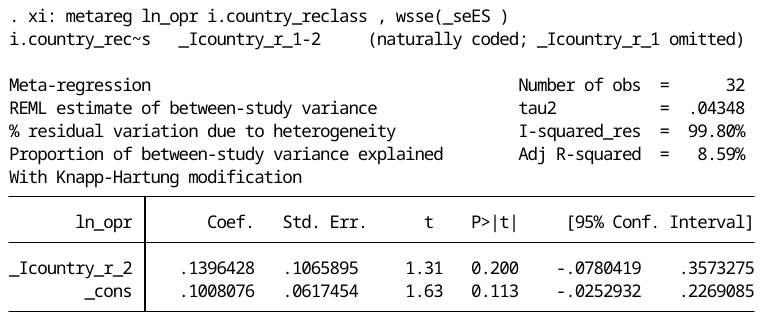

xi: metareg ln_opr i.continent , wsse(_seES )

None of the continents are significant – as p = 0.24 (for all covariates with Knapp-Hartung modification.

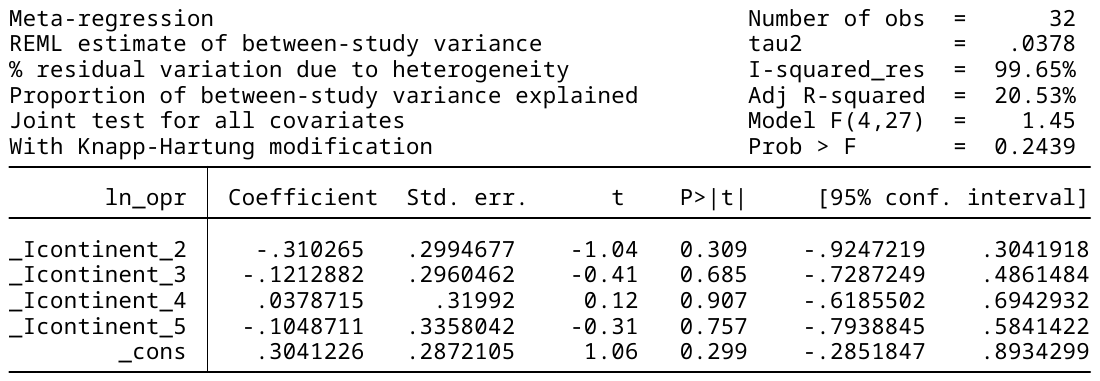

xi: metareg ln_opr i.study_type, wsse(_seES )

Study type (door-to-door survey vs predictive records linkage) does not explain heterogeneity p = 0.141 (metareg)

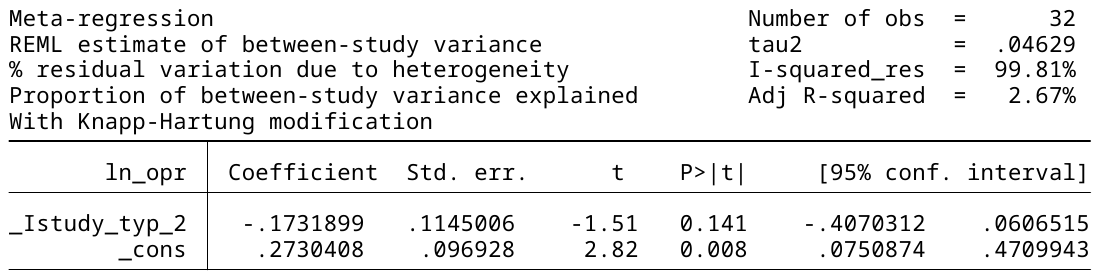

xi: metareg ln_opr i.publication_year , wsse(_seES )

Publication year does not explain data heterogeneity

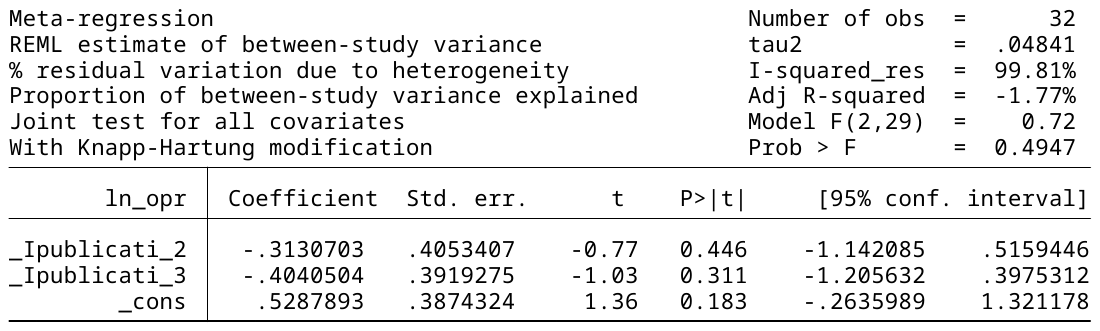

**#Run meta-regression for continuous variables:**

xi: metareg ln_opr f_to_m_difference_in_life_expect, wsse(_seES )

Female to male life expectancy difference in each country does not explain heterogeneity p = 0.080 (metareg)

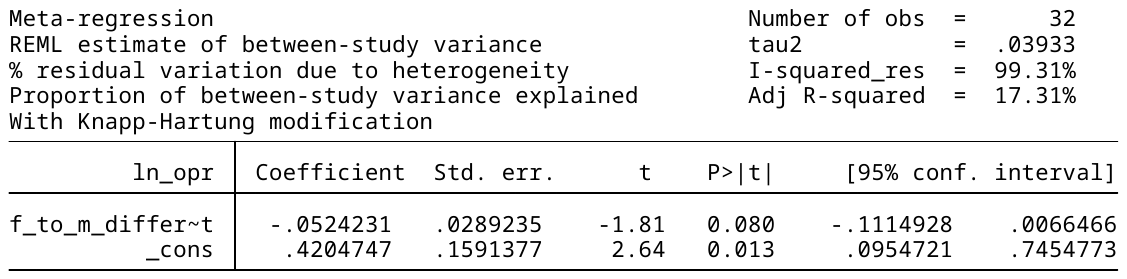

xi: metareg ln_opr median_age , wsse(_seES )

Median age does not explain heterogeneity p = 0.59 (metareg)

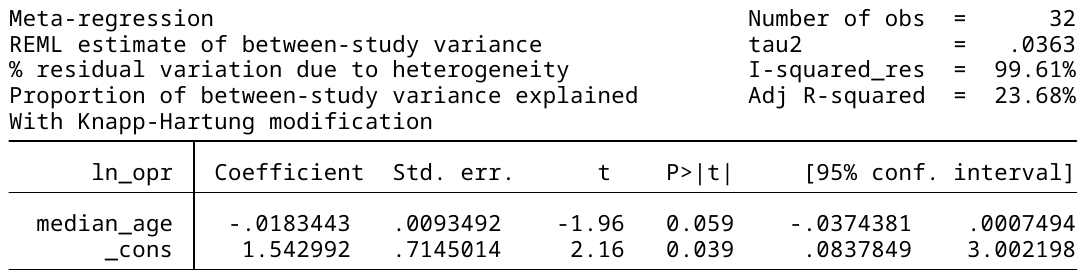

xi: metareg ln_opr age_life_exp_diff , wsse(_seES )

Difference between median age and life expectancy gap does not explain heterogeneity for prevalence (p = 0.288)

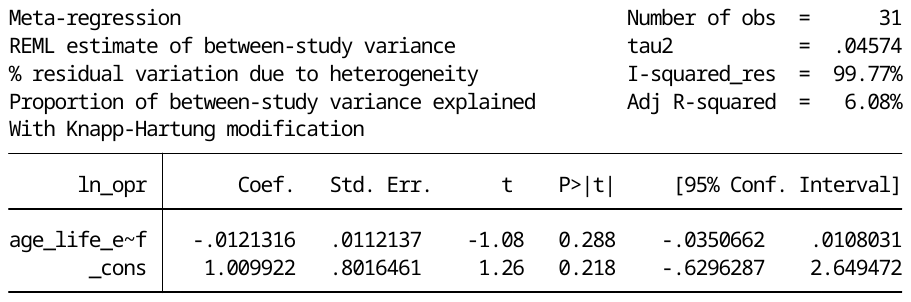

**#Run meta-analysis for subgroups – 1^st^ analysis:**

metan ln_opr ln_lower_ci_95 ln_upper_ci_95, random eform label(namevar= authoryear) by( country)

1 – HIC, 2 – L/MIC -> higher M/F ratio in HIC (1.258) than L/MIC (1.077).

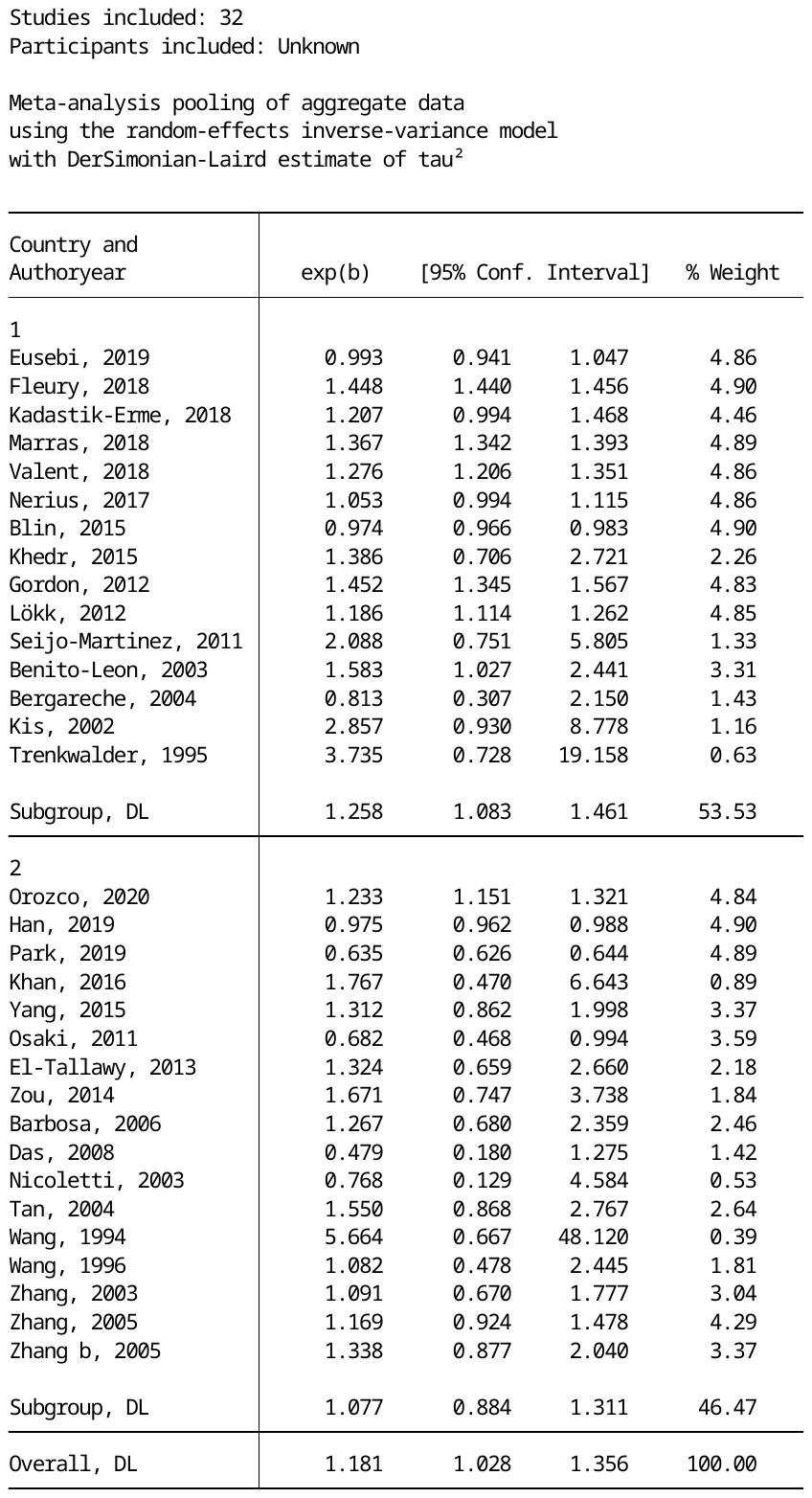

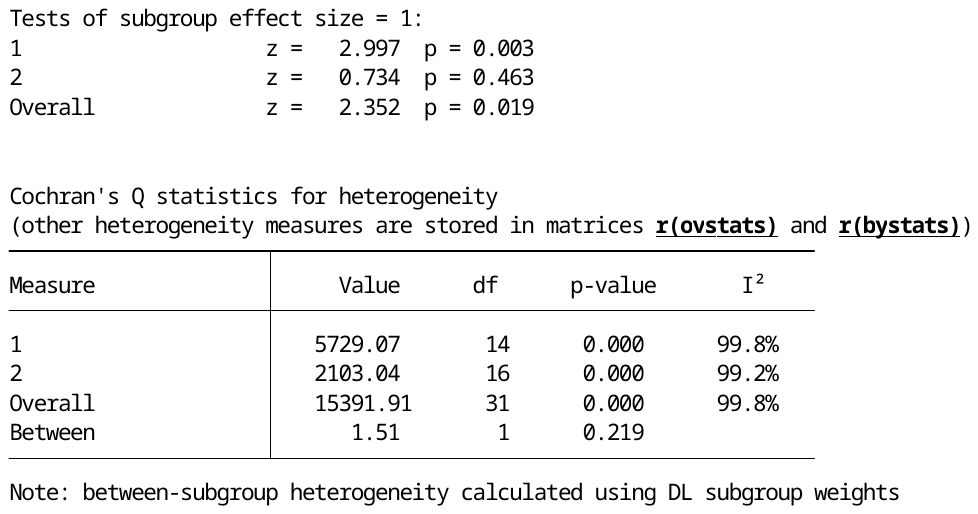

**Second analysis:**

metan ln_opr ln_lower_ci_95 ln_upper_ci_95, random eform label(namevar= authoryear) by( country_reclass )

1 – HIC, 2 – L/MIC -> lower M/F ratio in HIC (1.14) than L/MIC (1.3) – **opposite effect?**

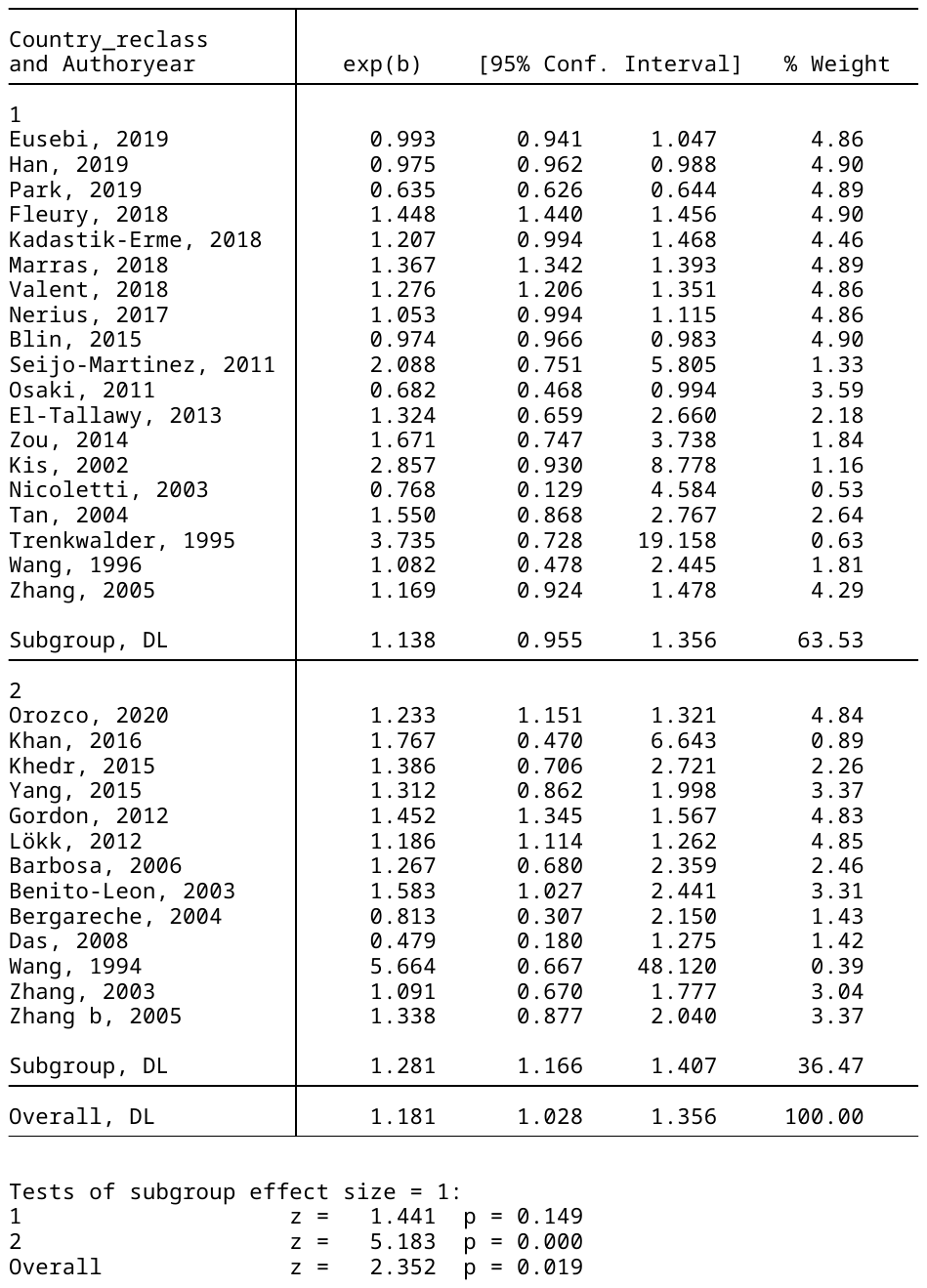

metan ln_opr ln_lower_ci_95 ln_upper_ci_95, random eform label(namevar= authoryear) by( co

> ntinent )

Asia lowest M/F ratio (1.037)

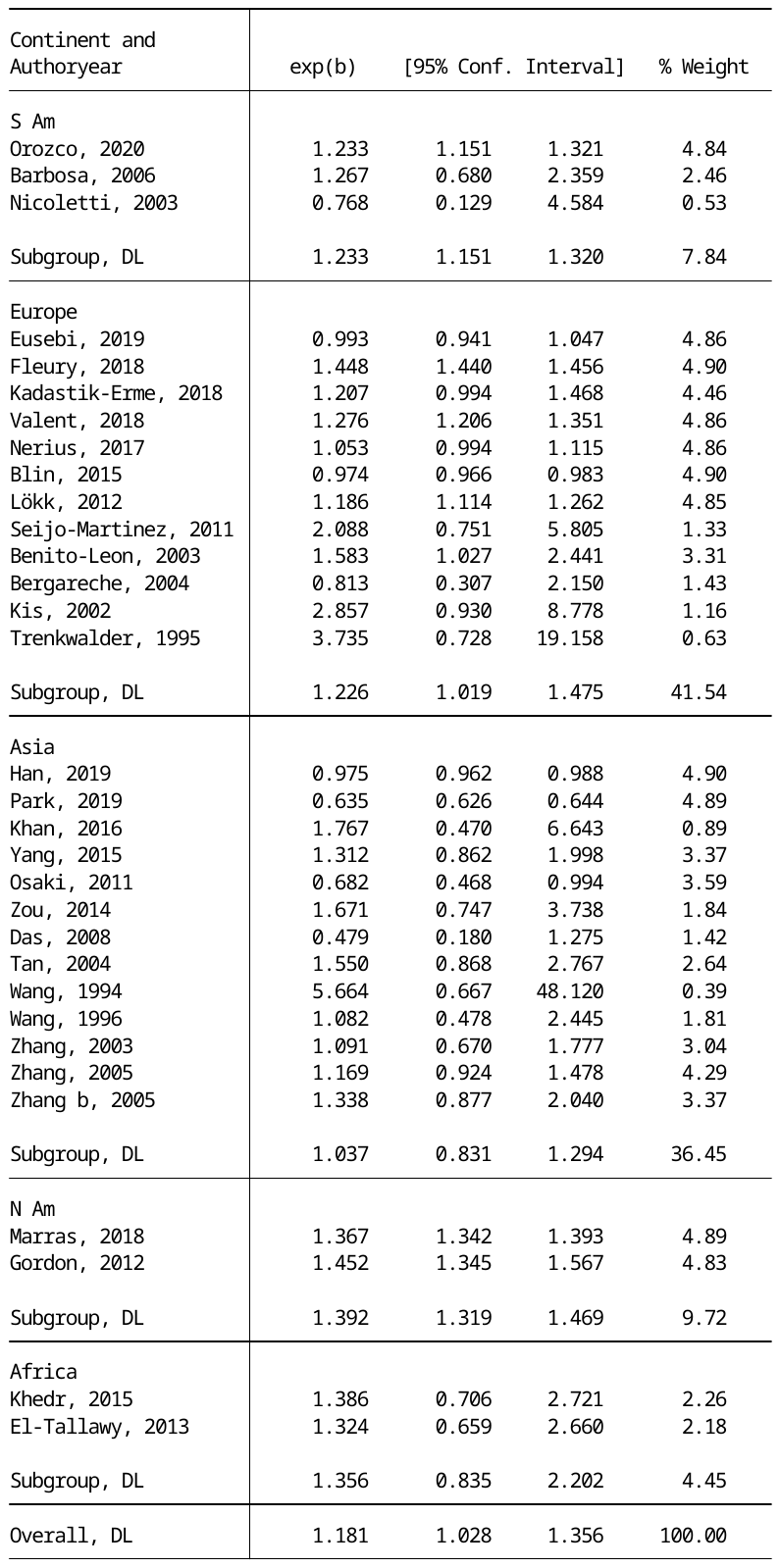

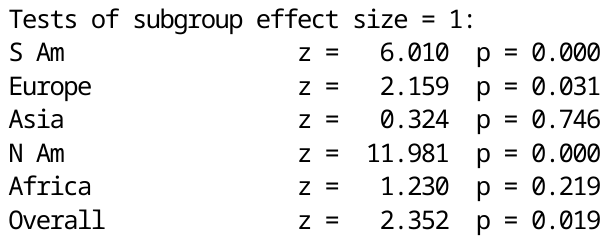

metan ln_opr ln_lower_ci_95 ln_upper_ci_95, random eform label(namevar= authoryear) by( publication_year )

M/F prevalence does decrease from 1990-2000 (2.06) to 2000-2010 (1.24) to 2010-present (1.15)

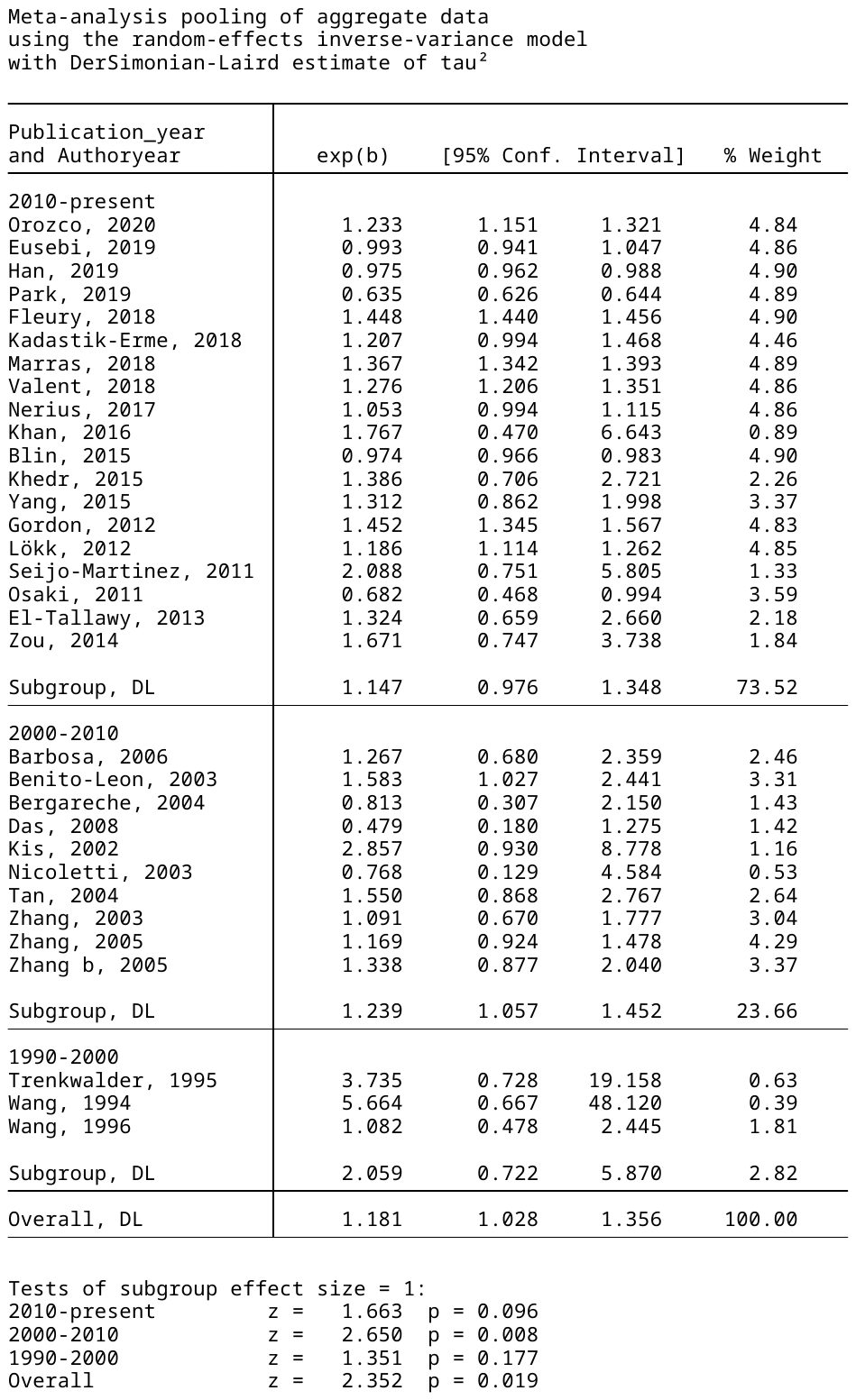

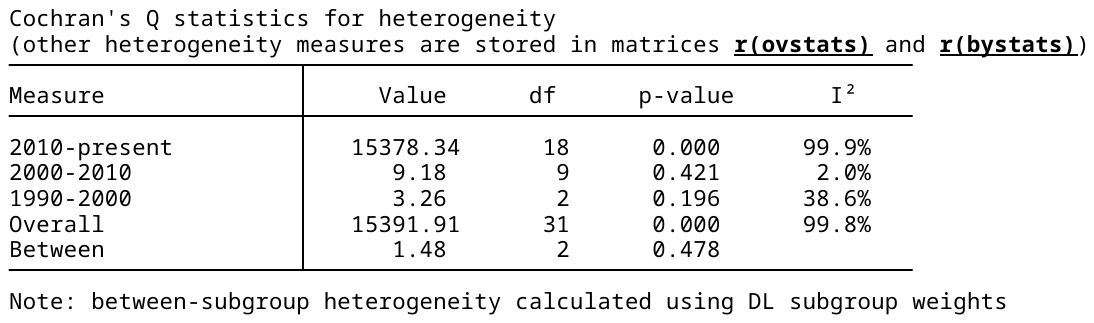

**metan ln_opr ln_lower_ci_95 ln_upper_ci_95, random eform label(namevar= authoryear) by(update)**

**Pringsheim 1.26; Us 1.15**

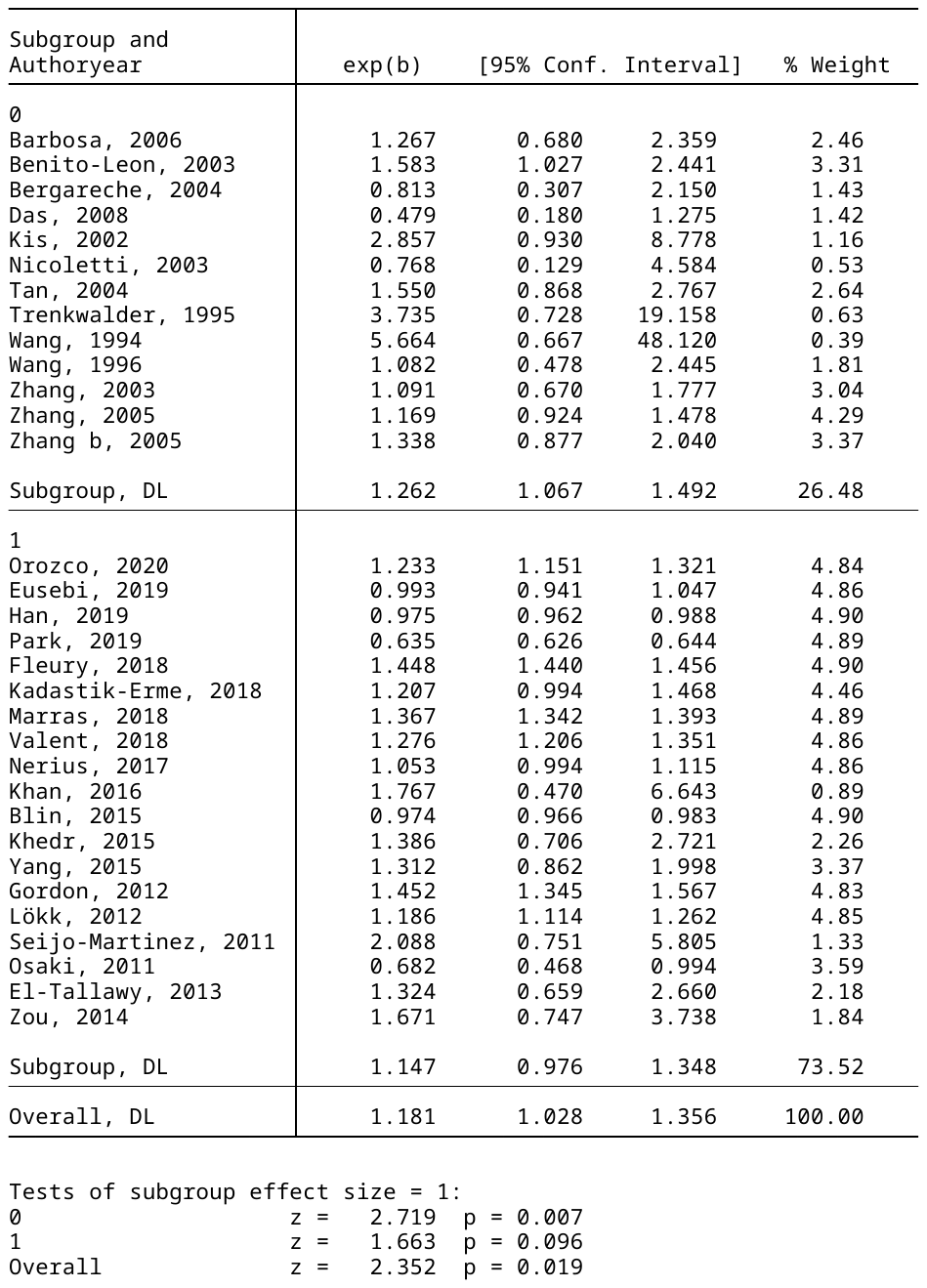

**INCIDENCE**

**#Set working directory and import csv file**

cd "H:\Gender MA\Data"

import delimited "H:\Gender MA\Data\ Incidence SR_AZ v2.csv"

**#Run MA:**

meta set ln_oir ln_lower_ci_95 ln_upper_ci_95, studylabel(author_year) random civartolerance(0.1)

metan ln_oir ln_lower_ci_95 ln_upper_ci_95, random eform label(namevar= author_year)

Overall RR 1.372 (1.22,1.53) ; Heterogeneity I^2: 98.2%

**#Run publication bias analysis:**

metabias ln_oir _seES, egger

metafunnel ln_oir _seES

There is no significant publication bias, p = 0.241 (metabias)

**#Run meta-regression for binary variables:**

First analysis

xi: metareg ln_oir i.country , wsse(_seES )

High vs low income does not explain the heterogeneity p = 0.017

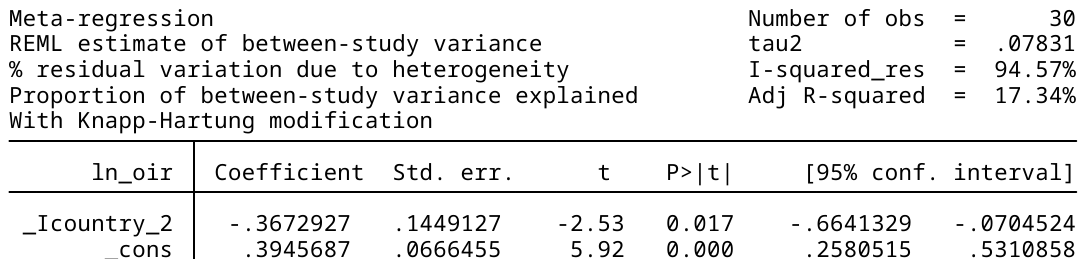

**Second analysis**

xi: metareg ln_oir i.country_reclass , wsse(_seES )

HIC vs non HIC as reclassified as per WB, not significant; p = 0.56

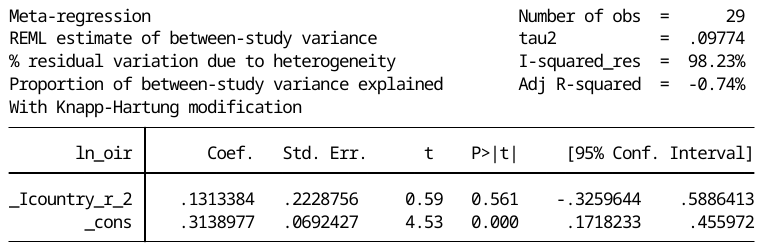

xi: metareg ln_oir i.study_type, wsse(_seES )

Study type (door-to-door survey vs predictive records linkage) does not explain heterogeneity p = 0.786

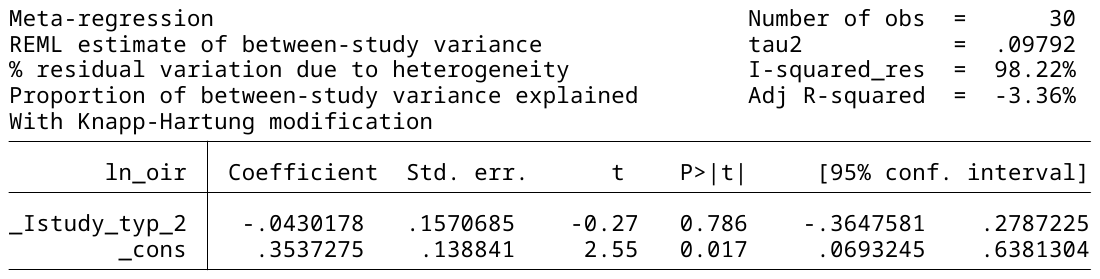

xi: metareg ln_oir i.Publication_year, wsse(_seES )

Publication year does not explain heterogeneity.

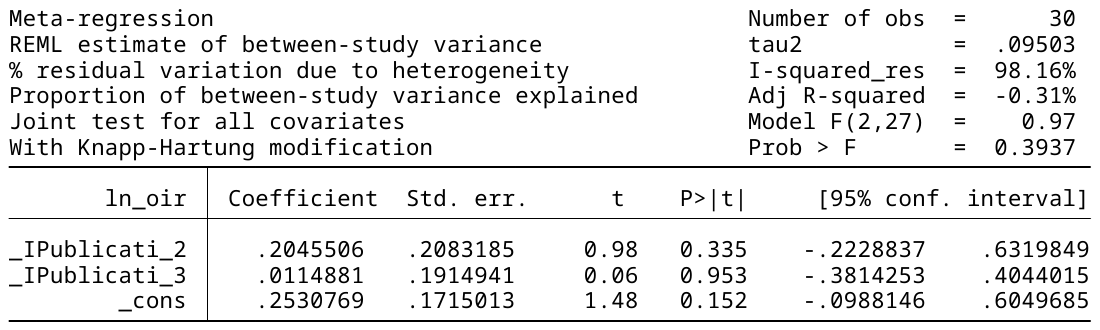

**xi: metareg ln_oir i.update, wsse(_seES )**

Validation, p=0.013 significant difference between the 9 new studies and the 21 old ones!! Bad.

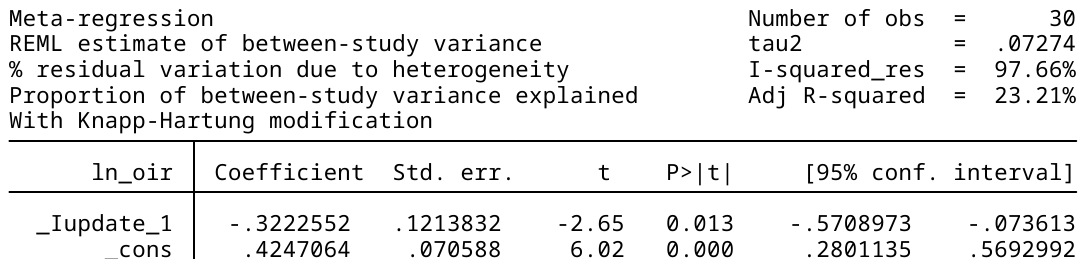

**#Run meta-regression for continuous variables:**

xi: metareg ln_oir f_to_m_difference_in_life_expect , wsse(_seES )

Difference in life expectancy does not explain heterogeneity for incidence -> p = 0.174

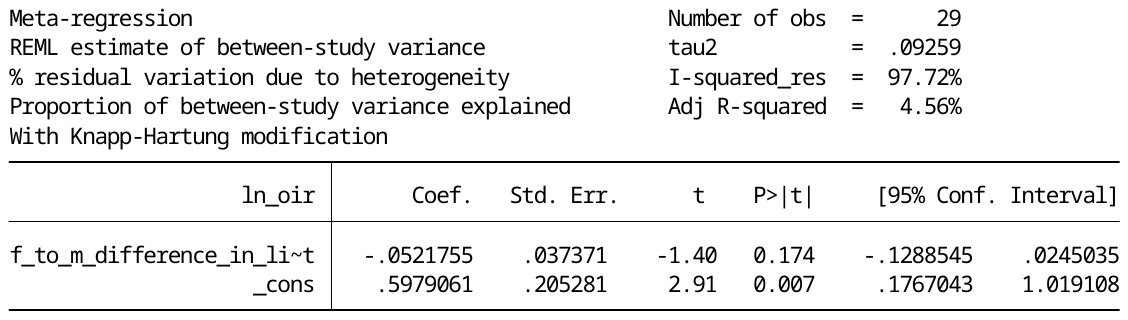

One study, from Taiwan, unable to get WHO data as it has been excluded from WHO in recent years (complex relationship to China).

F_to_M_difference_in_Life_expectancy

|  | F_to_M_difference_in_Life_expectancy |
| --- | --- |
| 1 | 4.85 |
| 2 | 6.66 |
| 3 | 6.66 |
| 4 | 4.15 |
| 5 | 4.85 |
| 6 | 4.76 |
| 7 | 4.53 |
| 8 | 6.38 |
| 9 | 4.01 |
| 10 | 4.02 |
| 11 | 4.85 |
| 12 | 6.49 |
| 13 | 5.82 |
| 14 | 5.05 |
| 15 | 3.75 |
| 17 | 3.25 |
| 18 | 3.75 |
| 19 | 3.75 |
| 20 | 3.75 |
| 21 | 5.74 |
| 22 | 3.76 |
| 23 | 3.74 |
| 24 | 6.78 |
| 25 | 5.05 |
| 26 | 6.31 |
| 27 | 4.65 |
| 28 | 3.75 |
| 29 | 4.53 |
| 30 | 11.78 |

!!!Without the last study from Russia (Winter 2010), it appeared that life expectancy did explain this heterogeneity (p=0.003). Why does one study skew things so much?

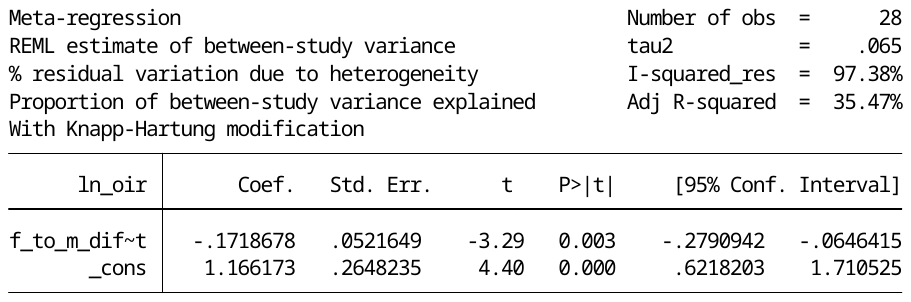

xi: metareg ln_oir median_age , wsse(_seES )

Median age does not explain heterogeneity (p = 0.276)

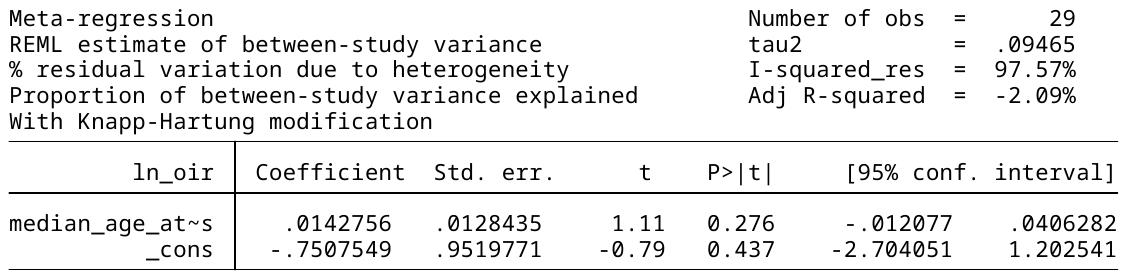

xi: metareg ln_oir Age_life_exp_diff , wsse(_seES )

Difference between age and life expectancy gap does not explain heterogeneity (p = 0.199)

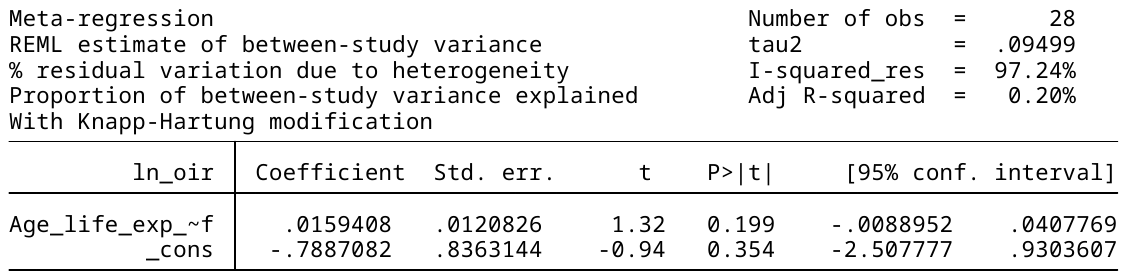

**#Run meta-analysis for subgroups**

First analysis

metan ln_oir ln_lower_ci_95 ln_upper_ci_95, random eform label(namevar= author_year) by(country)

1 – HIC ; 2 – L/MIC; Higher M/F ratio in HIC (1.473) vs L/MIC (1.003)

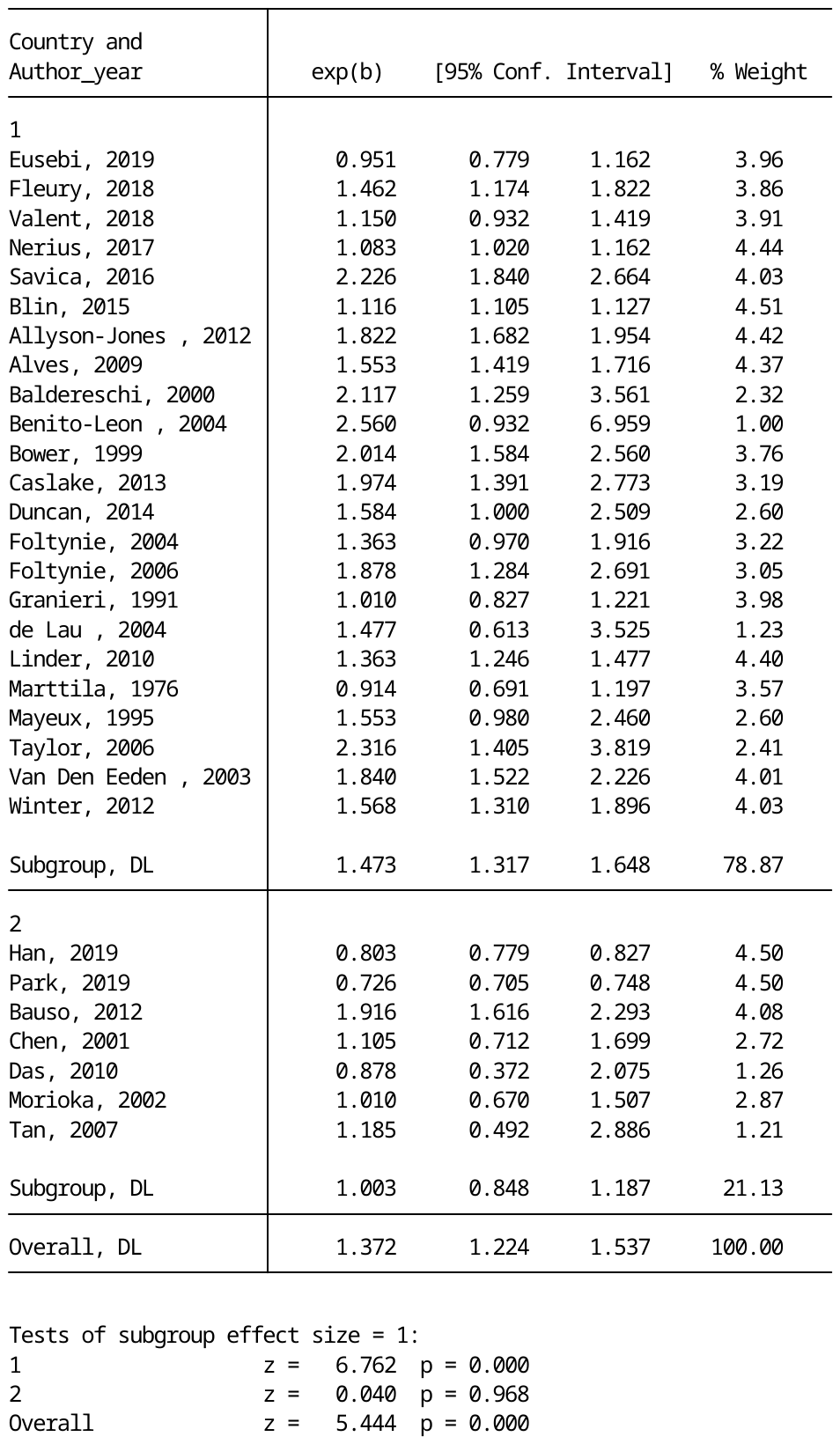

**Second analysis:**

metan ln_oir ln_lower_ci_95 ln_upper_ci_95, random eform label(namevar= author_year) by(country_reclass)

HIC 1.36; LIC 1.66

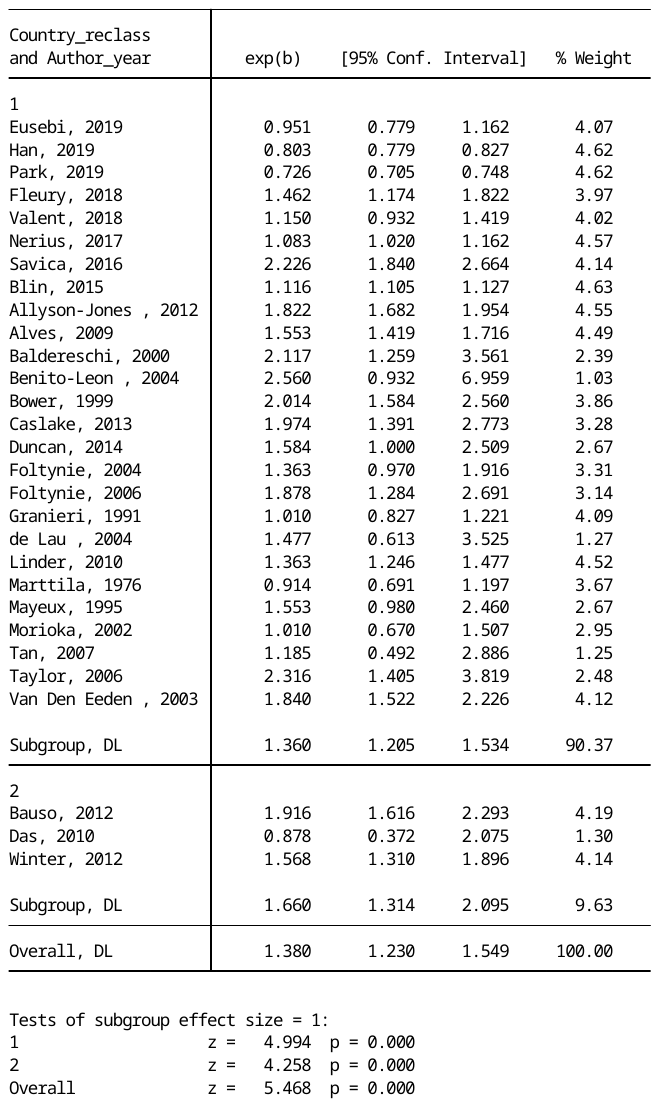

metan ln_oir ln_lower_ci_95 ln_upper_ci_95, random eform label(namevar= author_year) by(continent)

Asia lowest M/F ratio (0.792)

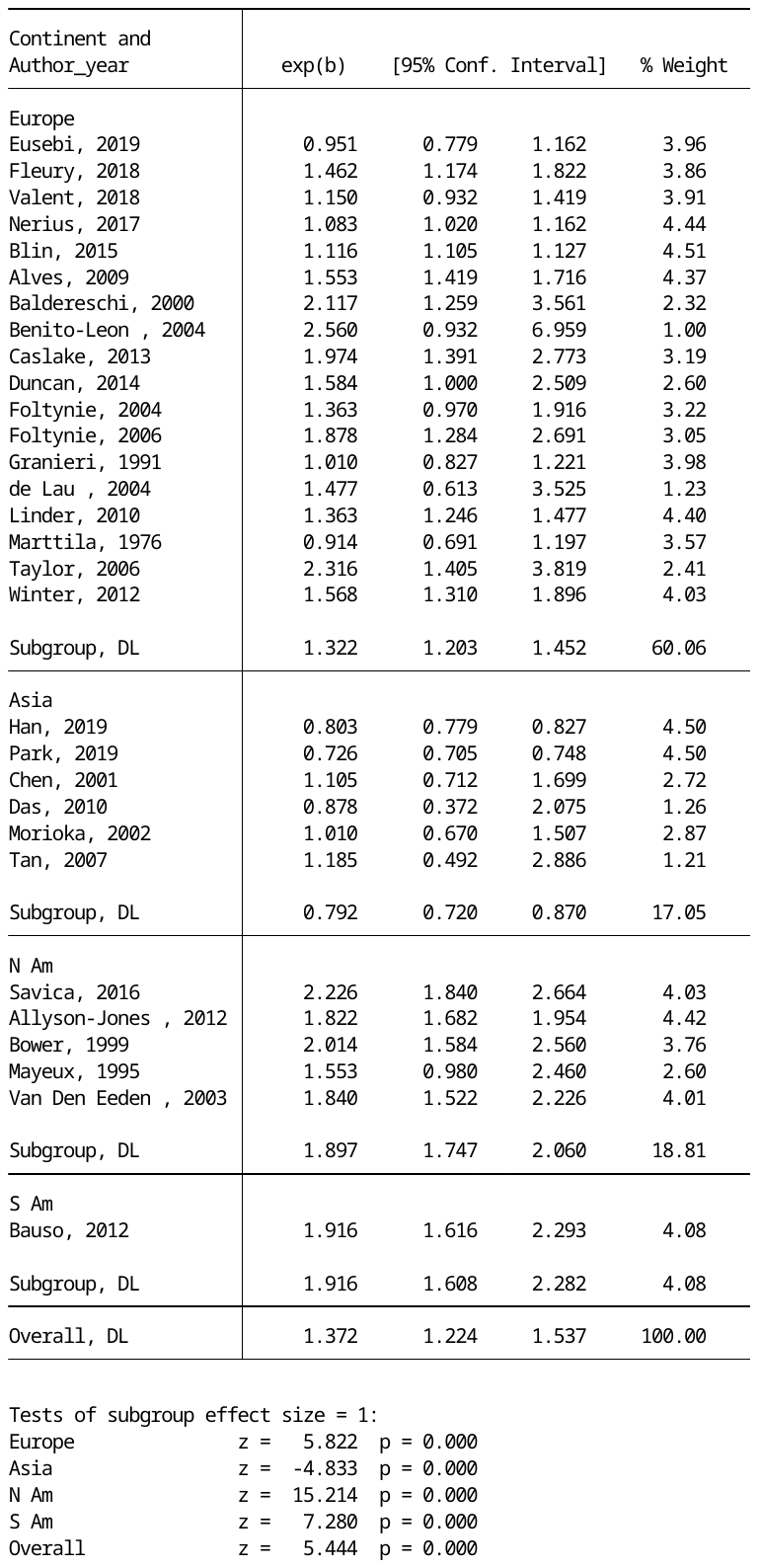

metan ln_oir ln_lower_ci_95 ln_upper_ci_95, random eform label(namevar= author_year) by(Publication_year)

1.3 -> 1.6 -> 1.37 (no trend)

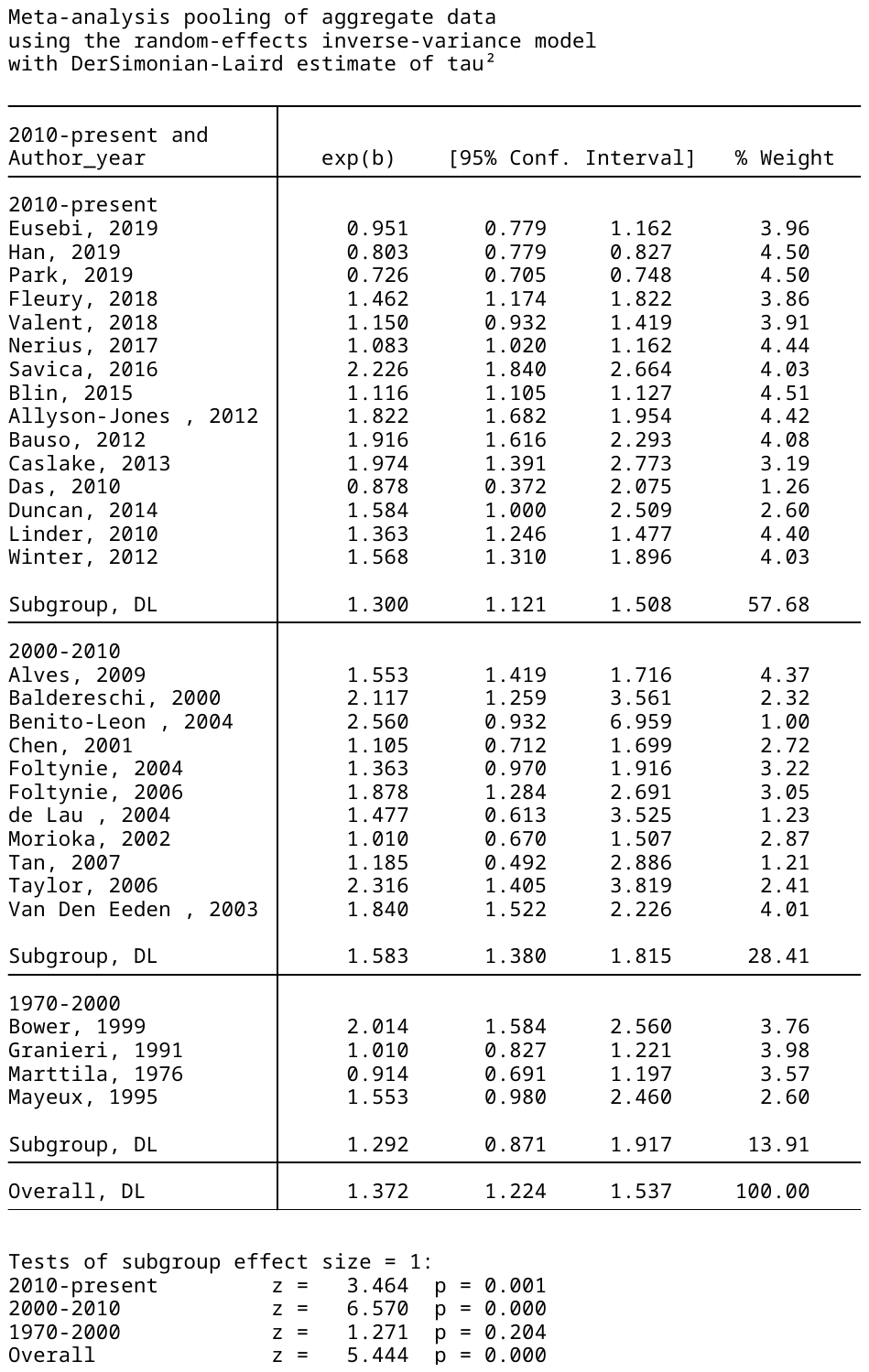

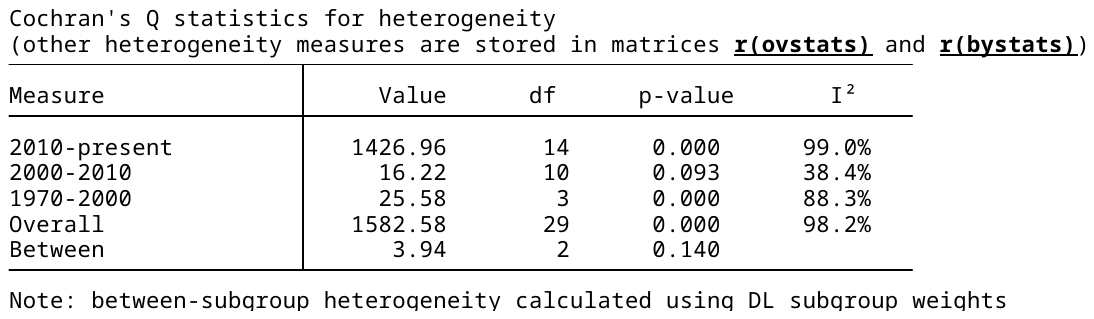

**metan ln_oir ln_lower_ci_95 ln_upper_ci_95, random eform label(namevar= author_year) by(update)**

Us 1.11 ; Moisan 1.53 – we have a lower ratio than Moisan **?why**

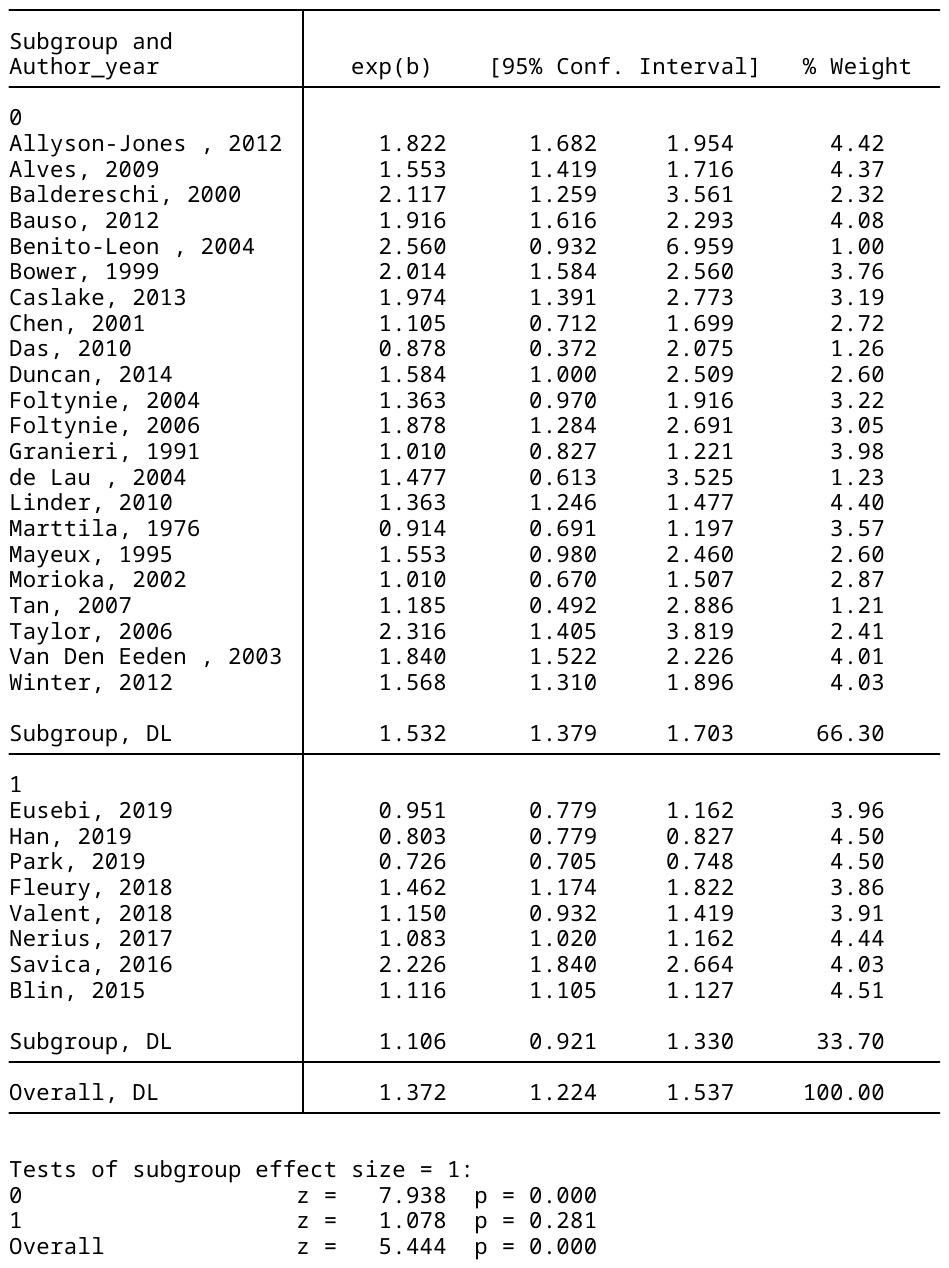
